# A phenomenological algorithm for short-range predictions of the Covid-19 pandemic 2020

**DOI:** 10.1101/2020.05.22.20098350

**Authors:** Piotr T. Chruściel, Sebastian J. Szybka

**Affiliations:** Faculty of Physics, University of Vienna; Astronomical Observatory, Jagiellonian University; Copernicus Center for Interdisciplinary Studies

## Abstract

We present an algorithm for dynamical fitting of a logistic curve to the Covid-19 epidemics data, with fit-parameters linearly evolving to the future. We show that the algorithm would have given reasonable short- and medium-range predictions for the mid-range evolution of the epidemics for several countries. We introduce the *double-logistic* curve, which provides a very good description of the epidemics data at any given time of the epidemics. We analyse the predictability properties of some naive models.

## 1 Introduction

There exist many sophisticated models trying to forecast the evolution of the Covid-19 pandemic. These require, for example, fitting of the epidemics data to data sets obtained by evolving differential equations with many parameters. Obtaining well motivated estimates for all these parameters is a difficult task, so guesses are unavoidable, leading to severe uncertainties concerning the predictive power of the models.

The question therefore arises, whether the epidemics data can be satisfactorily described by some explicit curves with a small number of parameters which are easily obtainable from the available data and, if so, whether such curves can be used to make predictions of the further evolution of the epidemics. The object of this work is to analyse the suitability towards this of a few empirical curves, to present an algorithm based on the logistic curve for short-term predictions of the epidemics, and to present the results of a study of the quality of this algorithm.

We note that most countries have infection curves with very similar shapes. There are some exceptions, with e.g. epidemiological curves for Denmark, Singapore or South Korea distinctively different from the remaining ones. A few other data curves (e.g. the Hubei province in China, France, Spain) have unusual jumps. Our predictability analysis only concerns typical curves, which are in vast majority.

## 2 Phenomenological curves

### 2.1 Logistic curve

Some authors have been using the *logistic curve* for describing the number *I*(*t*) of infected individuals. The function *I* is given by the equation

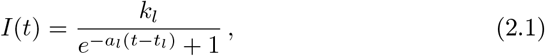

for some constants *k_l_*, *a_l_* and *t_l_*. Here the subscript *l* is a shorthand for “logistic”. The number *k_l_* is the predicted total number of infected individuals, the number *a_l_* governs the steepness of the curve, and *t_l_* is the inflection point of the curve, after which the daily number of new infected individuals starts decreasing. The curve (2.2) has the overall form seen in Figure 2.1.

**Figure 2.1:**
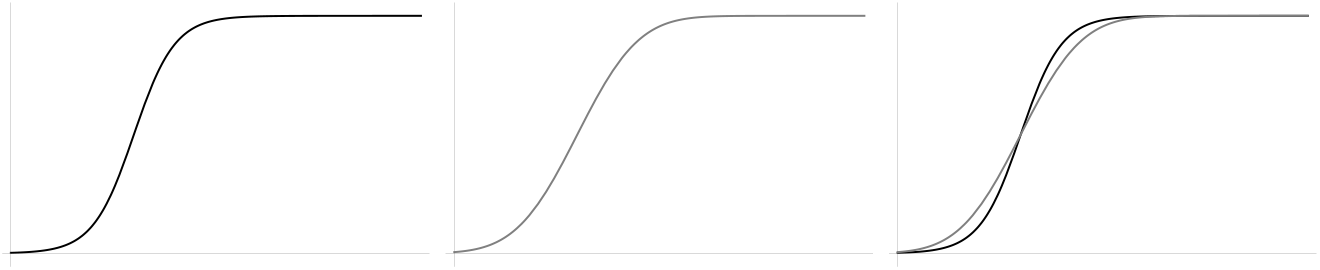
A typical logistic curve (left), Gauss-error curve (middle), and both curves superimposed (right).

The logistic curve provides a good description of many real-world phenomena, and is the exact solution to simple models of an epidemics [1]. Fitting the logistic curve to the Covid-19 data at any moment of time has been giving, as of the day of writing of this work, an optically satisfactory description for several countries, with decent confidence intervals for the parameters of the fits. We present some such fits in Figure 2.2. However, the fit parameters change in time so that fitting the logistic curve to the data has no value for long-range predictions of the epidemics.

**Figure 2.2:**
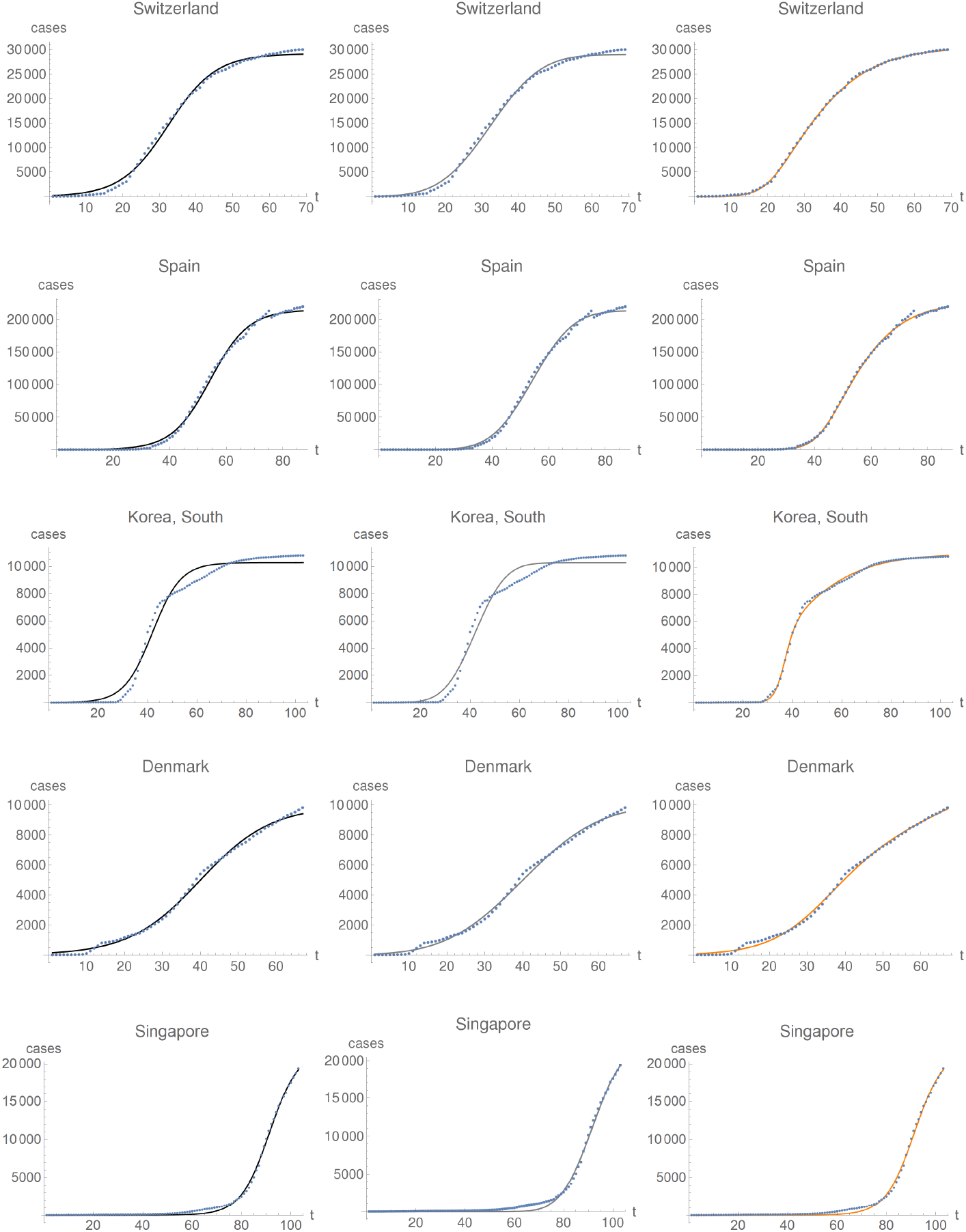
Fits of the logistic (left), Gauss-error (middle) and double-logistic curve (right) superposed with the data points on May 5 for selected countries. The days are counted from the day of beginning of the epidemics, which differs from country to country.

One can nevertheless try to test whether one can use the logistic curve for short-term predictions. We found that the resulting predictions typically undershoot the number of cases. For example, for Austria, in the (randomly chosen) period April 5 to May 5, 2020, next-day predictions were off-the-mark within the range between –5% to 0%, with more than two thirds of predictions between –4% and –3%; and one-week predictions were distributed between –10% and –5%, with about half of them between –6% and –5%. This is made precise by the histograms in Figure 2.3.

**Figure 2.3:**
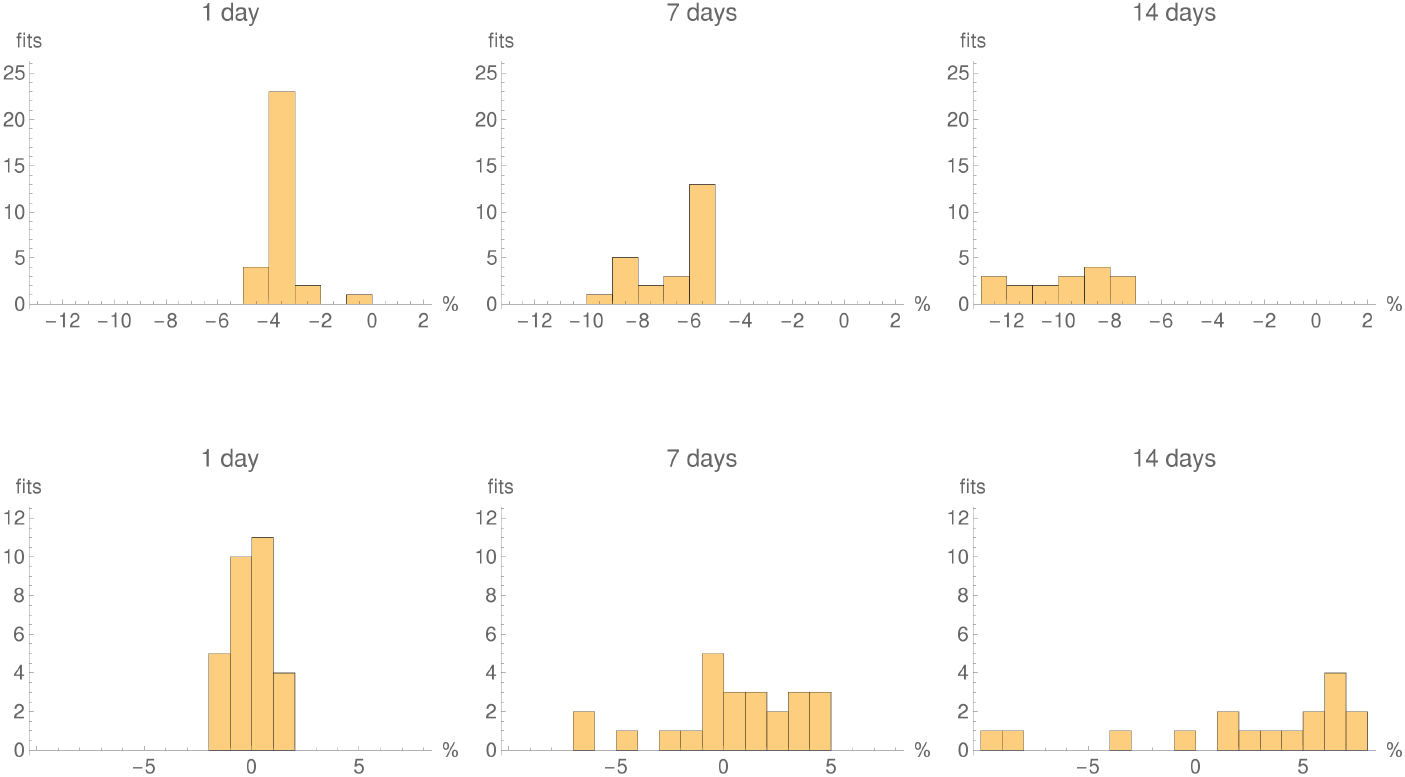
Histograms showing the number of predictions within 1% error bins, for Austria using the logistic fit (first row) and the double-logistic fit (second row), generated in the period April 5 - May 4, for 1 day, one week, and two weeks. Note a height rescaling between the first and the second row. The double-logistic fits are more scattered, but centred around 0%. The fact that the total number of fits on each histogram (here equal to the sum of the areas of the bins) decreases with the number of prediction days is due to the fact that we are testing predictions within a data set of prescribed total length.

This behaviour is typical for several countries.

This is clearly not satisfactory, and we will present a way of exploiting the systematic trends of these fits to propose an improved predictability algorithm shortly.

### 2.2 The Gauss-error curve

Other authors suggested to use the Gaussian-error curve, where *I*(*t*) is taken to be

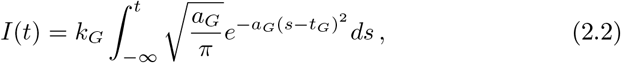

for some constants *k_G_*, *a_G_* and *t_G_*. Here the subscript *G* is a shorthand for “Gaussian”, the number *k_G_* is the predicted total number of infected individuals, the number *a_G_* governs the steepness of the curve, and *t_G_* is the inflection time of the curve. See Figure 2.1 for a typical plot.

Fitting the Gauss-error curve to the Covid-19 data at any moment of time gives again an optically satisfactory description for several countries, with decent confidence intervals for the parameters. In fact the fits look somewhat better than the logistic ones for many countries, whether by optical inspection of the curve (see Figure 2.2 for some examples), or by residuals, or by intervals of confidence of the parameters. But, similarly to the logistic curve, the parameters of the fit do not stabilise. Finally, at any day of the epidemics, the fitted numbers *k_l_* and *k_G_*, which provide a prediction for the total number of infected in their respective models, differ substantially for many countries.

Similarly to the logistic curve, this makes it clear that fitting the Gaussian-error curve to the data has no value for long-range predictions of the epidemics.

The short-term predictability properties of the logistic and the Gauss-error curves are very similar, with systematic undershooting of the number of new infections.

We use the built-in function `NonlinearModelFit` of Wolfram’s Mathematica to find our fits. It turns out that this function finds a fit to the logistic fit without difficulties, though with poor confidence intervals for the fitting parameters at the beginning of the epidemics. On the other hand, no fits at all are found for the Gauss-error function in early stages of the epidemics. This makes this curve much less useful in applications, if at all.

As a curiosity, we note that one can write down an ad-hoc system of SI equations for which *I*(*t*) is the Gauss-error curve:

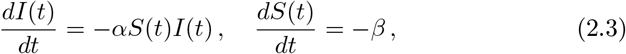

for some constants *α* and *β*. But it is not clear why the system (2.3) should describe well the evolution at hand globally, though perhaps confinement measures could produce effects modelled by the second equation in (2.3).

### 2.3 The double-logistic curve

In view of the above the question arises, whether some different families of explicit curves with a small number of parameters would fit better the problem at hand.

As such, by inspection of the majority of curves of total confirmed cases of Covid-19 the following picture emerges. The epidemics starts with the usual exponential growth, evolving into a forward-leaning-*S*-shaped curve without symmetry about the inflection point, and finishing in a very slow, optically linear dying-out phase.

We have made no attempt to include this last phase in our analysis, because for the period of data under consideration most countries have not reached a stable form of this last dying-out phase. In other words, we will be interested in a model where a good approximation of the end of the curve is a standard exponential dying-out of the epidemics.

Now, it is clear from the data that the initial exponential growth rate of the epidemics is different from the rate at which the epidemics dies out. On the other hand, both the logistic curve and the Gauss-error curve are symmetric in that respect. So better fits will certainly be obtained with curves which do not have this symmetry. An example of such a curve is obtained by taking a product of two logistic curves with different parameters. This leads us to consider the function

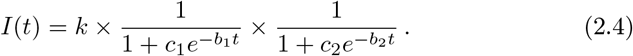

with five free positive parameters *k*, *b*_1_, *b*_2_, *c*_1_ and *c*_2_. For definiteness and without loss of generality we will assume that

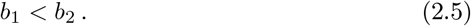

A simple analysis of (2.4) shows that:

1. The function *I*(*t*) tends to *k* as *t* tends to infinity, so that *k* describes the predicted total number of infected.
2. For large negative *t* the function behaves approximatively as

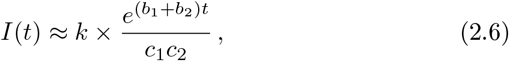 so that the initial rate of growth of the epidemics is *b*_1_ + *b*_2_, with growth coefficient *k*/(*c*_1_*c*_2_).
3. For large positive *t* we have

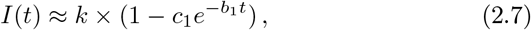 So that *b*_1_ describes the dying-out rate of the epidemics, with amplitude *kc*_1_

It is convenient to make trivial renaming of the parameters occurring in (2.4) to obtain, without loss of generality, the following form of the double-logistic function:

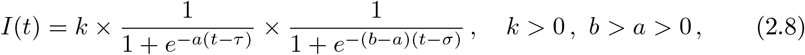

so that the parameter *a* has the interpretation of the dying-out rate of the epidemics, and *b* of the initial growth rate. Examples of plots of the curve (2.8) can be found in Figure 2.4.

**Figure 2.4:**
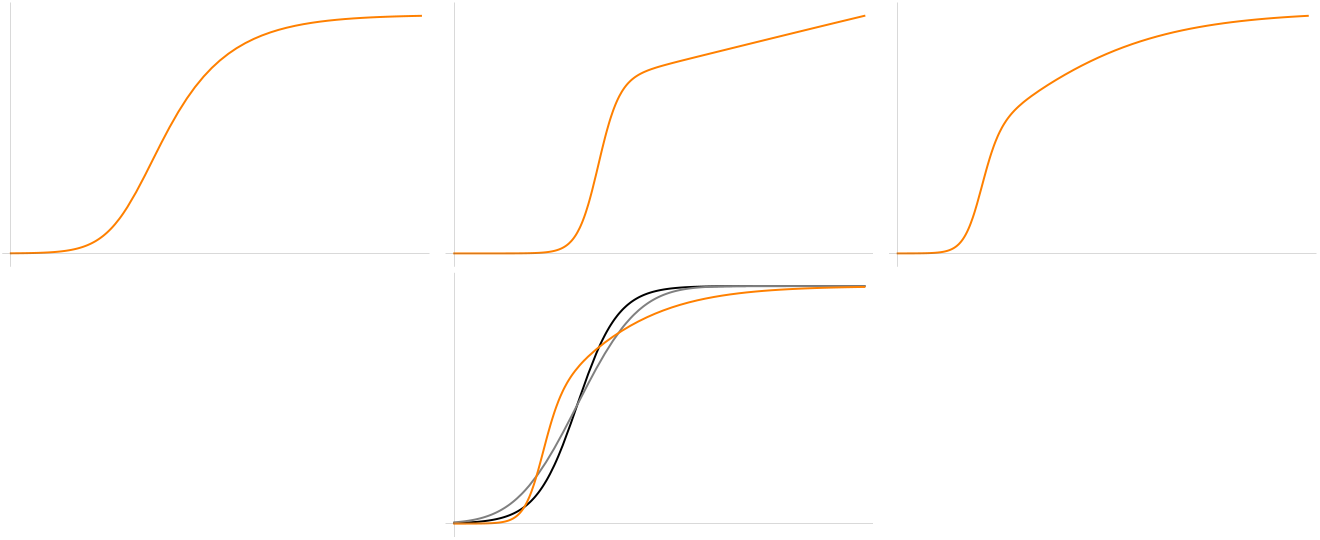
First line: examples of double-logistic curves. Second line: A superposition of a logistic (black) and a Gauss-error (gray) curve with *t_l_* = *t_G_* and same amplitude, together with a double-logistic (orange) curve with randomly-chosen parameters except for the total amplitude.

It is seen in Figure 2.2 that rather good daily fits to the Covid-19 data can be obtained using the function (2.4), although the fits are somewhat worse in the middle region for the wobbly time series of Denmark and Singapore. Compared to the fits previously discussed, the fits look much better optically; the residuals are smaller; the systematic undershooting at the end of the curve, inherent to the logistic and Gauss-error curves, disappears or is visibly reduced.

One finds, however, that the confidence intervals for the parameters are not as good as for the previous fits. Moreover, with more free parameters Mathematica’s fitting procedure is very unstable and sometimes requires a trial-and-error procedure to find a fit, if at all.

On one hand, the fitting parameters do not stabilise in time either so that this curve is again unsuitable for global predictions. On the other, the short-term predictions are typically better than for the previous curves, *if* stable fits are available. For example for Austria, all one-day predictions in the period April 5- May 5, 2020, were within 2% of the data, with about 2/3 of them within 1%; four day predictions within 5% of the data with about half within 1%; and one week predictions in an interval of 7% around the observed value, with more than one third within 1%. This can be seen in Figure 2.3.

While this is better than the short-time predictions based on the logistic curve, we will see in Section 3 how to obtain much better such predictions.

Ignoring its relatively poor predictability properties, the double-logistic curve seems to provide a good description of the overall features of the confirmed-cases curves and deceased curves for many countries. This could make this curve useful for further comparative studies of the Covid-19 epidemics.

We finish this section by noting that the *Richards curve* [2], which has been used in various models of population dynamics, is somewhat related to our double-logistic curve. We comment on this in Appendix B.

## 3 The algorithm

Let us present an algorithm which would have produced more accurate short-term predictions of the dynamics of the epidemics, as compared to the ones discussed above, for many countries, when used in the period April 5 - May, 5, 2020. The dates here are chosen for definiteness, and in a random manner.

The algorithm proceeds as follows.

We denote by {*n*_1_, *n*_2_, …} the time-series of the number of confirmed infected individuals for a given country. In what follows we will often simply say “infected individuals”, meaning the number of *confirmed infected cases* as available on the John Hopkins University (JHU) server.

Let us write the logistic curve (2.2) as

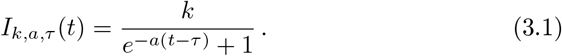

We denote by *k_i_*, *a_i_*, *τ_i_*, the values of the parameters *k*, *a*, and *τ* of the curve (3.1), determined by fitting the data for the epidemics curve from the start of the epidemics to day *i*. For conciseness of notation we write

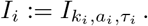

Thus, for *ℓ* ≤ *i* the number *I_i_*(*ℓ*) is the number of infected at day *ℓ*, as approximated using the best fit, calculated at day *i*, to the infection data known up to day *i*. For *ℓ* > i the number *I_i_*(*ℓ*) would then be the prediction of the model to the future.

If the curve predicted correctly the development of the epidemics, the numbers *k_i_*, *a_i_* and *τ_i_* would remain constant, within their confidence intervals, as *i* changes. It turns out that this is not the case: in the period covered by this analysis, for several countries the parameters so obtained change approximately linearly with *i*. In order to compensate for this,

1. we make a linear fit to these “running constants”, at time *i*. For *j* > 0 this provides us an *extrapolation to the future* of the values of (*k*, *a*, *τ*), which we denote by

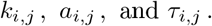 Thus the values *k_i_*, *a_i_*, and *τ_i_* are known from the data at day *i*, while for *j* > 0 the values *k_i,j_*, *a_i,j_*, and *τ_i_*,*_j_* are calculated by linear regression from data known up to day *i*.
2. We calculate the shift, say *s_i_*, so that *I_i_*(*i*) + *s_i_* coincides with the observed data on day *i*:

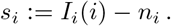 This compensates for the systematic undershooting of the logistic curve.
3. Finally, for *j* > 0 the number

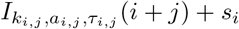 is the prediction, on day *i*, of the number of confirmed infections on day *i* + *j*.

We have applied this procedure to several countries using the data available from the JHU server. The results of the algorithm, as applied to Austria, are presented in Section 4. A sample of countries is similarly analysed in Appendix A. A systematic simultaneous study for several countries is presented in Section 6.

We refer to the above method of making predictions as *the logistic fit with running constants*.

The algorithm is quite successful when tested on the epidemic curve of Italy. There, again in the period April 5 to May 5, 2020, all one-day predictions were within 1% of the data, not exceeding 3% for eight days, all but one within 3% for thirteen days (the outlier within 4%), with a maximal error less than 4% for twenty-nine days and 5% for thirty days. This can be read-off from Figure A.5.

As explained in more detail below, the way we implement the above requires at least 18 data points, a number which has been chosen randomly. The point is that in order to gain any confidence in the usefulness of the procedure one has to have a sufficient collection of reasonable logistic fits. This is not the case at the beginning of the epidemics, where a simple exponential provides instead a good description of the dynamics.

Our attempt to apply an analogous procedure based instead on the double-logistic curve failed to produce successful results. We believe that this is due to the numerical instability of the fits obtained, as was clear in some of our attempts to do so. It is conceivable that the double-logistic curve together with a more sophisticated fitting algorithm could provide better predictability results than our algorithm based on the straightforward logistic curve, but this remains to be seen.

Alternatively, since the Gauss-error function also produces better fits than the logistic curve, it would be natural to use our algorithm using the Gauss-error function. We did not do this because the fits to that last function seem to be less stable, when using Mathematica, compared the logistic ones. In particular Mathematica finds no fit at all to the Gauss-error function in the early stages of the epidemics, while fits to the logistic curve are found.

## 4 The fits

We present here some more details of the analysis discussed above.

All data are retrieved from the John Hopkins University (JHU) server, URL http://raw.githubusercontent.com/CSSEGISandData/COVID-19/master/csse_covid_19_data/csse_covid_19_time_series/time_series_covid19_confirmed_global.csv

The analysis presented here is based on the cumulative data set downloaded from the JHU server on May 6. This data set contains data up-to-and-including May 5, 2020. We note that data at the JHU are sometimes corrected backwards in time, so making fits on data sets downloaded on a later day and truncated to May 5 could lead to different results for the countries where backwards changes have been made.

Now, many data sets at JHU start with a long string of zeros. We disregard those zeros and, unless explicitly indicated otherwise, day one of the epidemics on all our plots is the first day at which there are at least two cases of confirmed infected individuals.

Some time series in the JHU data set contain repeated entries, which are unlikely in the middle of a fast spreading epidemics. We interpret this as a weekend effect, or an accidental omission by the reporting authorities, who made up for this on the next day by inputting the same cumulated number of new infections twice. In order to smooth-out the resulting jumps in the curve we replace systematically the first point of such a pair by the average of neighbouring points. No other alterations to the time series have been performed. We have not attempted to study whether further smoothing of the data would result in better predictability properties of our algorithm.

All fits are done using the `NonlinearModelFit` procedure in Mathematica. In order to find a good fit, or a fit at all, the procedure sometimes requires providing by hand the values of the parameters near which the fit should be performed. Most fits have been found automatically, by using the number of infected on the day of the fit as the initial value of the parameter *k* for the fitting procedure, with the starting points of the remaining parameters set to average values observed (roughly) in our analysis so far, independently of the country. For the logistic fit, the starting value of *a* for the fitting procedure was 0.08 and that for *τ* was 22, unless indicated otherwise by hand for unstable fits. In order to steer the system away from poor fits we imposed that the search for a fit should be done with the constraints that 0 < *a* < 2 and that k be smaller than 100 times the number of infected cases at the last day of the fit; these values being compatible, by a huge margin, with all good fits we have inspected.

It is appropriate to provide more details concerning the logistic fit with running parameters.

First, as already pointed out, we decided randomly that the epidemics starts when there are at least two infected individuals. This is counted as day one of the epidemics, all previous zeros and ones in the time series are ignored.

Next, we decided randomly that no time series which, after the above adjustment, are shorter than 18 days will be considered. The procedure is then somewhat different according to whether the series is shorter or longer than 25 days.

Suppose that the time series is between 18 and 24 days. We then calculate the best fit parameters to the logistic curve ending at the last eight days of the series. Hence, every fit to a logistic curve is based on at least eleven data points. Using these eight logistic curves, we make four linear fits to the values of each of the parameters *k*, *a* and *τ* of (3.1), one for each of the four intervals containing 8, 7, 6 or 5 successive days which are anchored at the last day of the time series. This is done using standard linear regression. For each of the parameters *k*, *a* and *τ* we choose the best linear fit, amongst the four, to extrapolate that parameter to the future. The method is illustrated in Figure 4.1.

**Figure 4.1:**
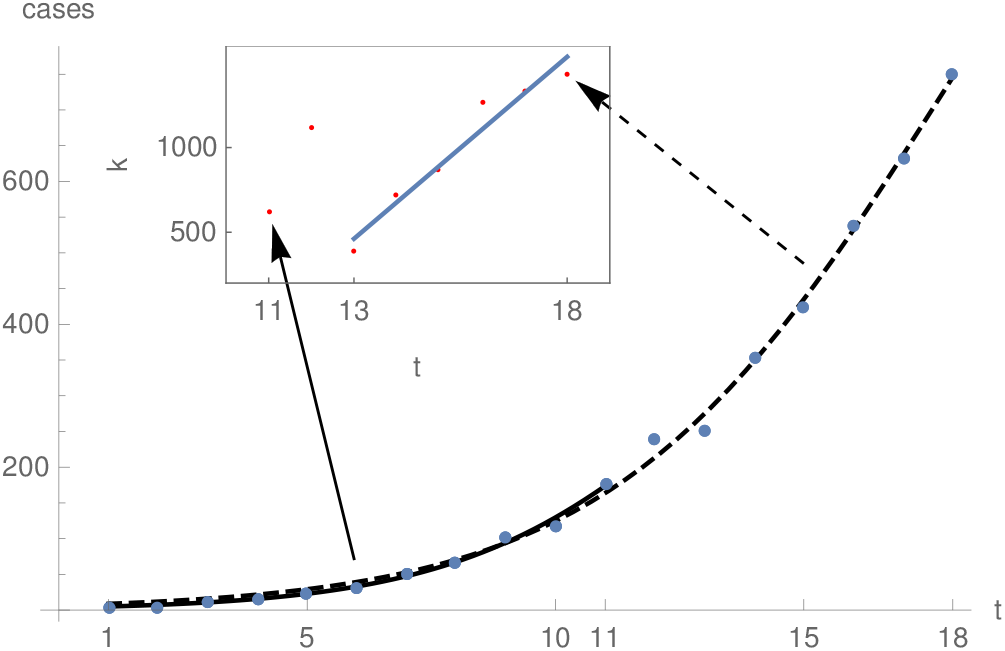
The figure illustrates the algorithm for a time series with 18 data points. The first logistic fit, plotted with a continuous line, based on the first eleven data points, gives the first value of *k*, indicated by a red point on the inset plot. The next value of *k* is obtained using a fit to the first twelve data points, and so on, until the fit based on all eighteen data points, plotted by the dashed curve, which provides the last value of *k*. The system finds the best linear fit for the values of *k*, with last point anchored at the last value of *k* (as obtained from the dashed logistic curve). This gives the blue line in the inset plot; in this example the best linear fit is based on the six last values of *k*.

If the time series is longer than 24 days, the procedure is identical except that we seek the best linear fit for the 11 intervals ending at the last day of the time series, with the length of the intervals varying from 14 to 4.

All the numbers above have been chosen randomly, and we have not attempted to study whether some other choice would lead to an algorithm with better predictability properties.

## 5 Austria

We continue with the details of our analysis on the example of Austria. The results of a similar analysis for some selected time series from JHU can be found in Appendix A.

In Figure 5.1 we show the latest fits to the double logistic, Gauss-error, and logistic curve, including the residuals (defined as the difference between the data and the fitting function) and the fit parameters. We note that a good fit should lead to residuals without any obvious structure. So, from this point of view, none of the fits is satisfactory, with the double-logistic fit providing the optically-best residuals in any case.

**Figure 5.1:**
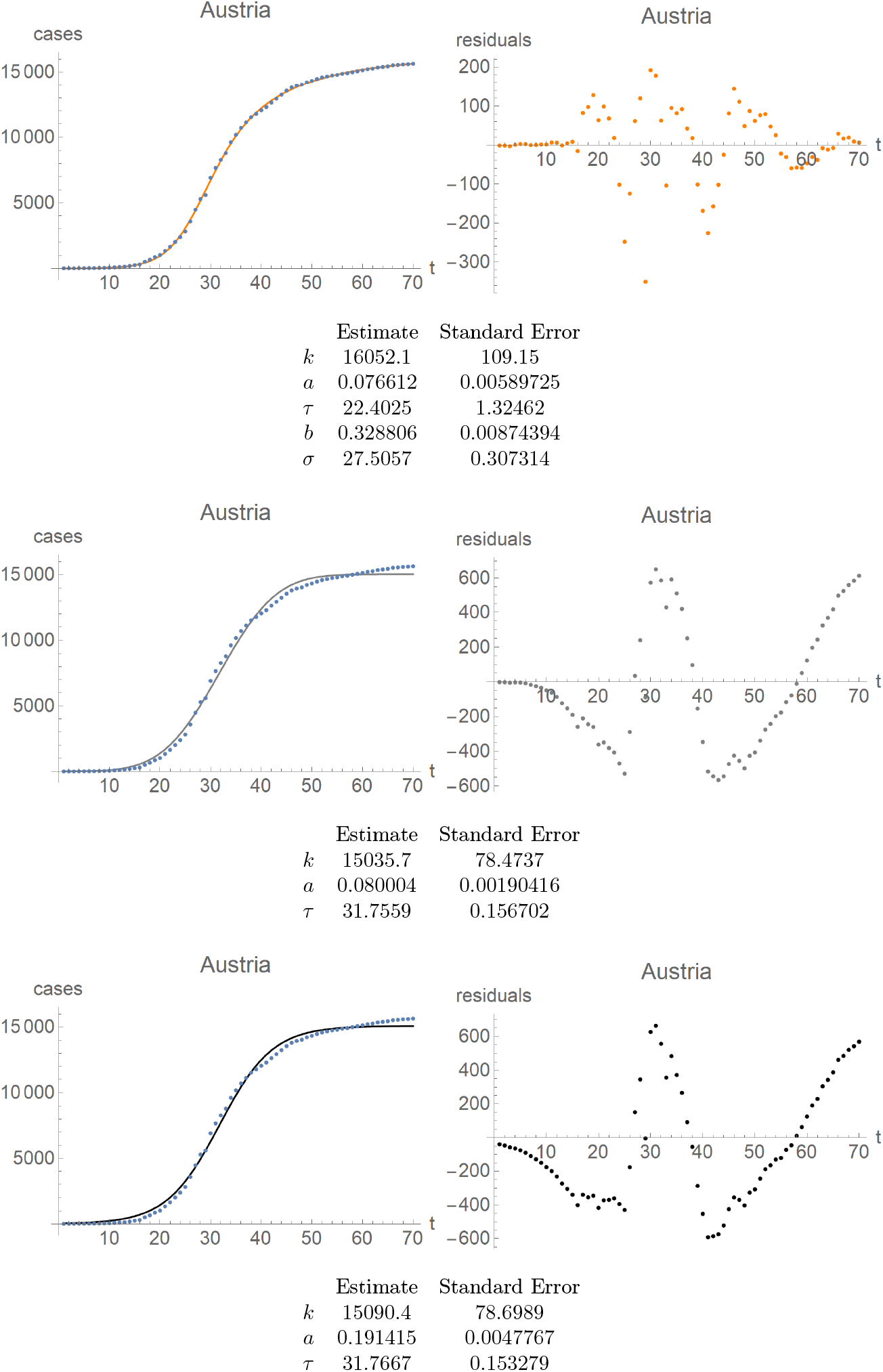
Austria. First row: fit of the double-logistic curve (left) and its residuals (right). Same for the Gauss-error curve in the second row, and the logistic curves in the third row. Note distinct scales for the residual plots as compared to the fitted curve, as well as a distinct scale for the residuals of the double-logistic curve compared to the other residuals.

In Figure 5.2 we show how the parameters of the fits, and their confidence intervals, change in time, for the double logistic, Gauss-error, and logistic curve.

**Figure 5.2:**
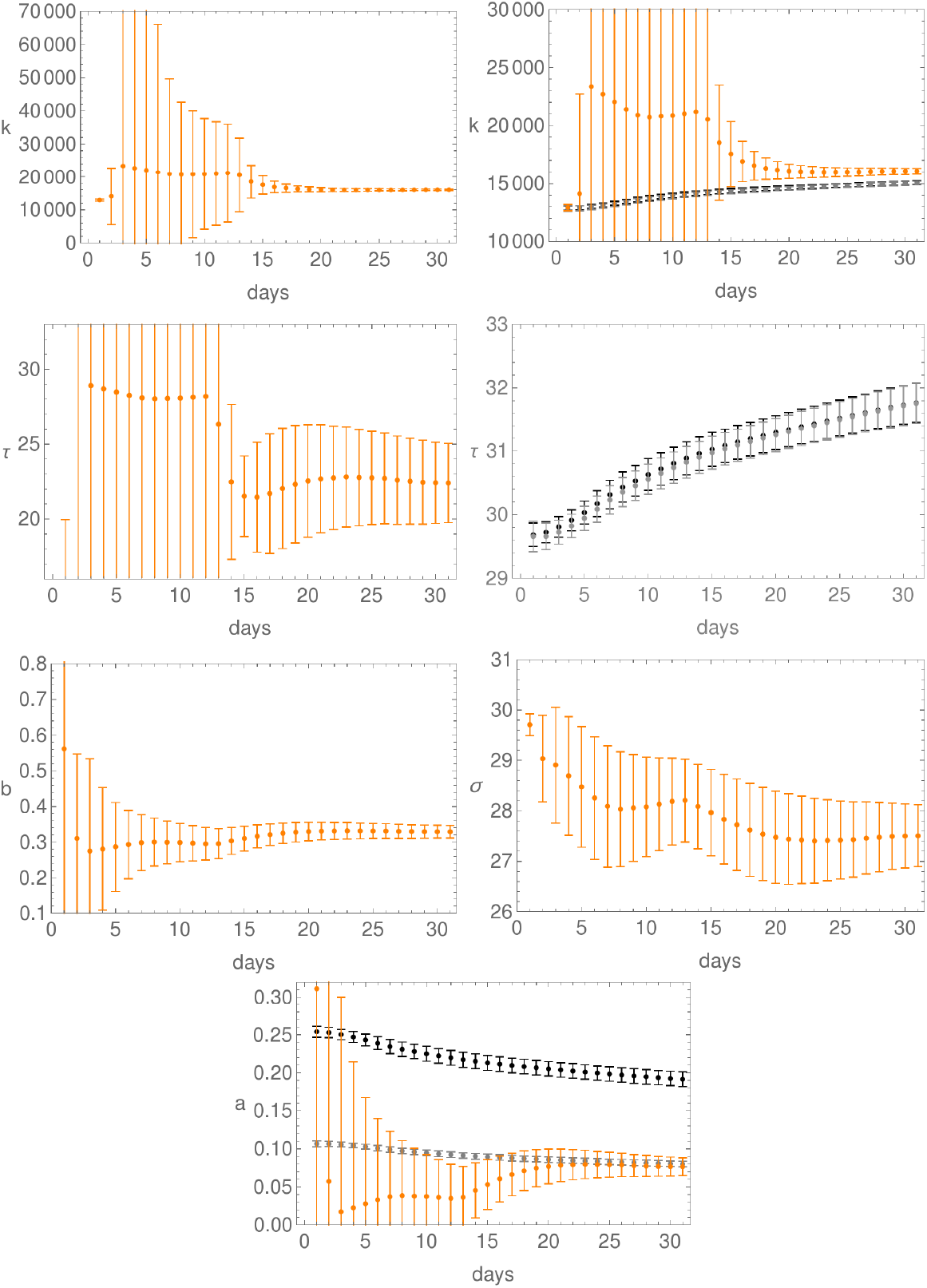
Evolution of the fit parameters in time, together with their confidence intervals, for Austria; day 1 is April 5. First row: parameter *k* for the double-logistic curve (left), and for all three curves (right). Here, as elsewhere, orange is for the double-logistic, black for the logistic, and grey for the Gauss-error function. Second row: the *τ*-parameter for the double-logistic curve (left) and for both the logistic and Gauss-error curves (right). Third row: plots of the parameters *b* (left) and *σ* (right) for the double-logistic curve. Last row: plot of the parameter *a* for all three curves.

In order to test the predictability value of our algorithm we proceed as follows. For each day in the period April 5 - May 4 we calculate the number of confirmed infected individuals, as predicted by our algorithm, for the remaining days of the period. We then calculate, for each day, the difference between the predicted number with the observed number, as provided by the JHU server. We express this difference as a percentage of the observed number. This gives us a string of relative differences between the observed number and predicted one, including sign: a positive sign means that our algorithm overshoots, a negative one means that the algorithm undershoots.^1^

We will refer to these sequences of predictions as “predictability strings”. Thus, the string starting on April 5 contains 30 numbers between -100 and 100, the first entry in the string being the relative error of the one-day prediction for April 6, the last entry being the relative error for the thirty-days prediction for May 5. We emphasise that this only uses data points known on and before April 5. The string starting on April 6 will contain 29 numbers, with the last prediction for May 5, 30 days ahead. The last “string”, generated on May 4, will consist of one number only, the relative error of the one-day prediction for May 5.

All the resulting strings for Austria are plotted in the first row of Figure 5.3. Such plots will be referred to as “predictability plots”.

**Figure 5.3:**
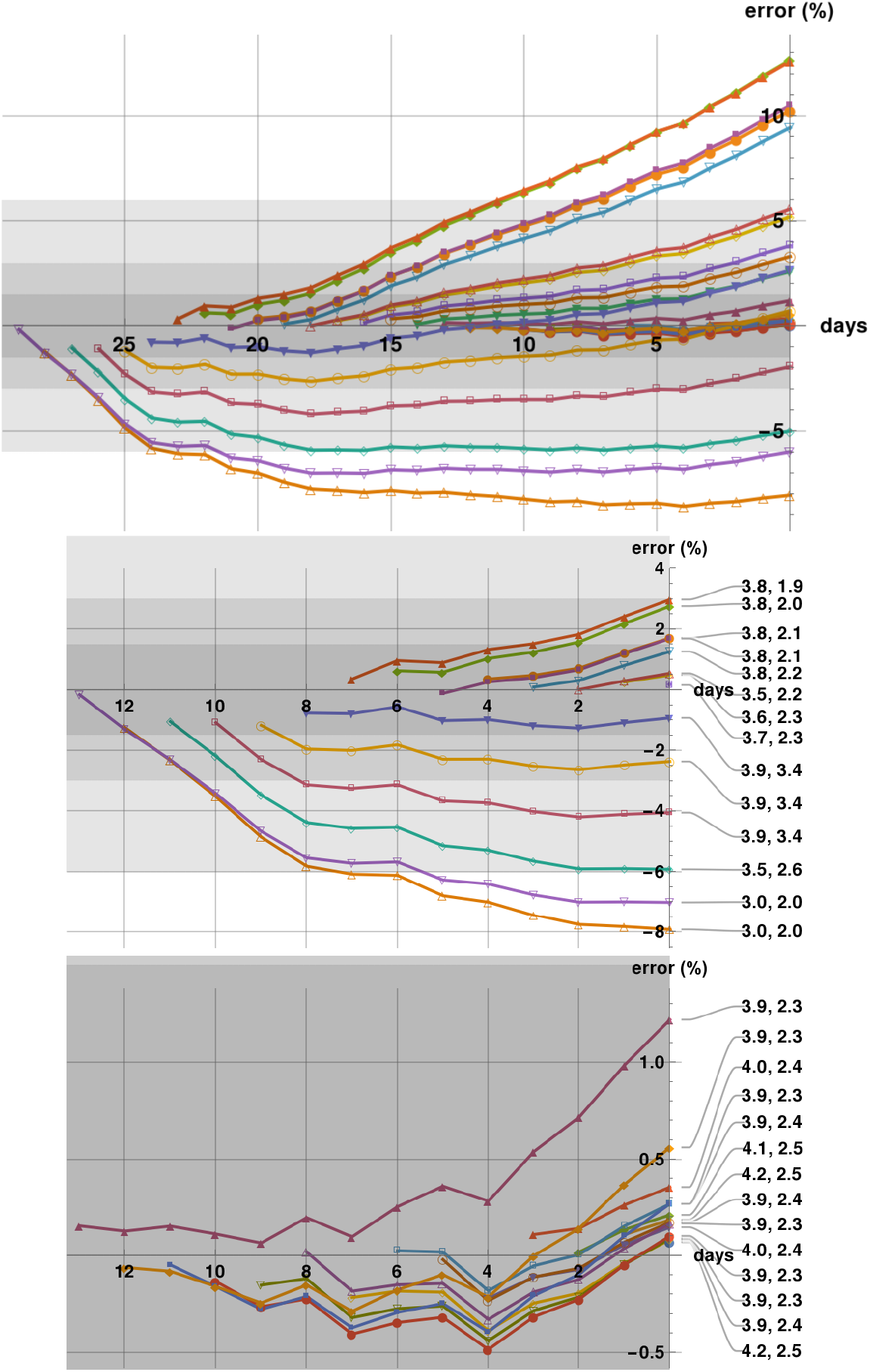
Predictability plots for Austria generated in the period April 5 - May 4. The first row shows all predictability strings with last prediction on May 5, which is the last day on the first and third rows. The shadings correspond to a 1.5%, 3%, and 6% error zones. The second row shows the predictability strings cut-off at April 20, with badness parameters indicated. The third row shows only these predictability strings which start within a two-weeks period ending on May 5. Almost all fits for Austria have comparable, relatively small first badness parameter, but this is not the case for countries with wobbly time series. It can be seen that fits with larger second badness parameter have worse predictability properties.

The predictability strings generated above will contain 30 one-day predictions, 29 two-days predictions, 23 seven-days predictions, etc., with only one 30-days prediction. In Figure 5.4 we show the histograms of the errors of the predictions so obtained using one-percent bins. We see that one-day predictions in the period April 5- May 5, 2020, were within 1.5% of the data, with more than 4/5 of them within 1%; four day predictions within 5% of the data with more than half within 1%; and one week predictions in an interval of 7% around the observed value, with more than one third within 1%.

**Figure 5.4:**
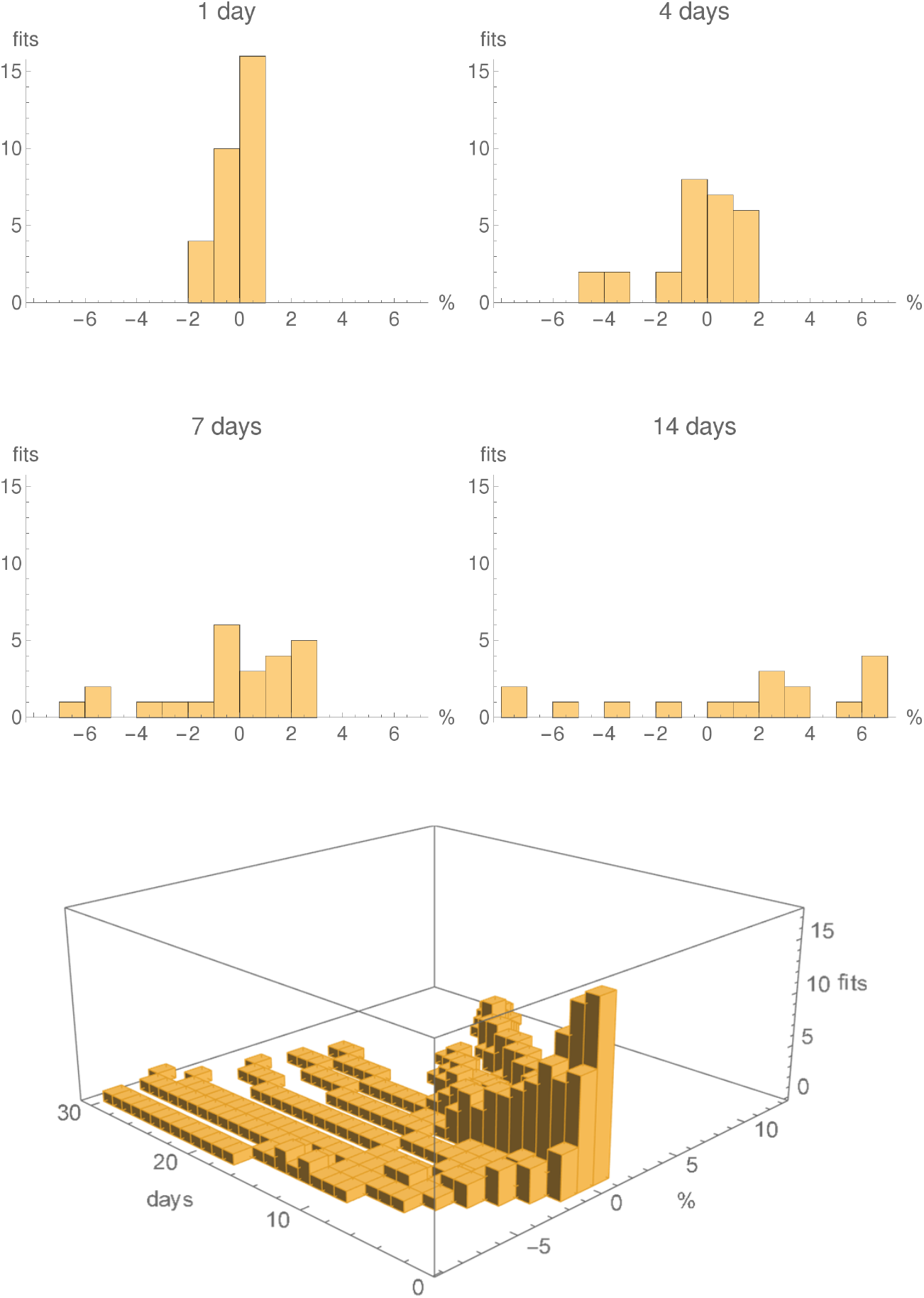
Histograms showing the numbers predictions within 1% error bins, for the logistic fits with running parameters, for Austria, generated in the period April 5 - May 4, for 1 day, 4 days, one week and two weeks. The last row shows a 3D histogram for all 30 fits. The individual histograms in the first two rows are slices, at the given number of days, of the 3D histogram.

Not unexpectedly, the later in the epidemic, the better the predictability of the algorithm. This can be read directly from the first row Figure 5.3, but is seen more clearly when comparing the second and third rows in that figure. The third row of Figure 5.3 shows only those predictions which start on April 21 or later. There we also included information on the “badness” of the predictability strings, to be explained in detail shortly; roughly speaking, this is a pair of numbers, the first being the maximal error, in percents, of the fits used to generate the predictability string, with the second the maximal relative error, again in percents, of the parameter *k* of the fits used to generate the predictability string. The third row of Figure 5.3 should be compared to the second one, where the predictability strings starting on April 5 are truncated to the two-weeks period ending on April 20.

Inspection of the predictability plots shows that some fits fare significantly worse than most. The question then arises, whether one can filter such fits out in advance, to obtain better predictability results. The idea is to associate to each fit “badness numbers”, which could be used to predict how bad is a fit likely to be. For instance, on some days an automatic search for a double-logistic fit, or a fit to a Gauss-error curve, did not succeed. In such cases Mathematica sometimes returns zero, or returns what looks like a curve with a random set of parameters, or returns a curve using the suggested input parameters. Clearly no useful prediction to the future of such a data point can be done. The result of the fit comes with a warning from the system, in which case an ad-hoc subjective intervention by hand can sometimes solve the problem. But this is not practical for an automatic procedure for the statistical analysis of many thousands of predictability data carried out in Section 6. So we ignore the warnings and let the system calculate the associated predictability strings, but we need a method to exclude such results from the predictions.

For this we implemented the following procedure. To each fitted curve we associate a pair of “badness numbers”. The first badness number is calculated as the size of the absolute values of the largest residual in the fit divided by the value of the fit at the end of the fitting period, multiplied by 100. Since all the curves under considerations are monotonically increasing, the badness number measures thus the size, in percents, of the absolute value of the largest residual, relatively to the largest number in the current fit.

Here a few comments are in order:

1. A perfect fit would have zero first badness number, and no such fits have been observed for obvious reasons.
2. In all our calculations a fit had first badness number 100 if and only if Mathematica returned the zero function as a fit, which happened sometimes when the fitting procedure did not succeed.
3. The first badness number ignores the fact that the fit can be very poor in the initial period of the epidemics, where the numbers are much lower than the number of cases at the end of the fitting period. This feature is not a problem from our perspective, since we are interested in extrapolating the epidemics curve to the future, hence in the region where the numbers are large. So a fit can have a low badness number when it fits well the data towards the end of the epidemics even though the fit is very poor at the beginning.

Next, in order to calculate our predictions, we need reliable values of the parameters (*k*, *a*, *τ*) of the fit. Our second badness number for a logistic fit is the maximum value of the relative standard errors of the parameters *k*, a and *τ* in percents; in other words, it is the maximum of a) the standard error of *k* divided by the estimate of *k* and multiplied by 100, and b) the standard error of a divided by the estimate of *a* and multiplied by 100, and c) the standard error of *τ* divided by the estimate of *τ* and multiplied by 100. We note that there is a lot of freedom of optimising this, and we made no attempts to do this.

We emphasise that the badness parameters are known at the date of the fit, and that it is clear from the predictability tables in Appendix A that they can be used *a priori* to filter-out the blatantly wrong predictions by setting up thresholds. These thresholds need to be fine-tuned individually for each country.

The badness parameters of the logistic fit with running parameters are set to be the maximum value of the badness parameters of those logistic fits which have been used to determine the best linear interpolation of the parameters. For example, if four logistic fits have been used to provide the linear interpolation of the parameter *k*, and three for the parameter *a*, then the maximal values of the badness parameters of the four relevant fits are deemed to be the badness parameters of the fit.

Plots of the first badness parameter can be found in Figure 5.5, and plots of errors of *k*, *a* and *τ* in Figure 5.6.

**Figure 5.5:**
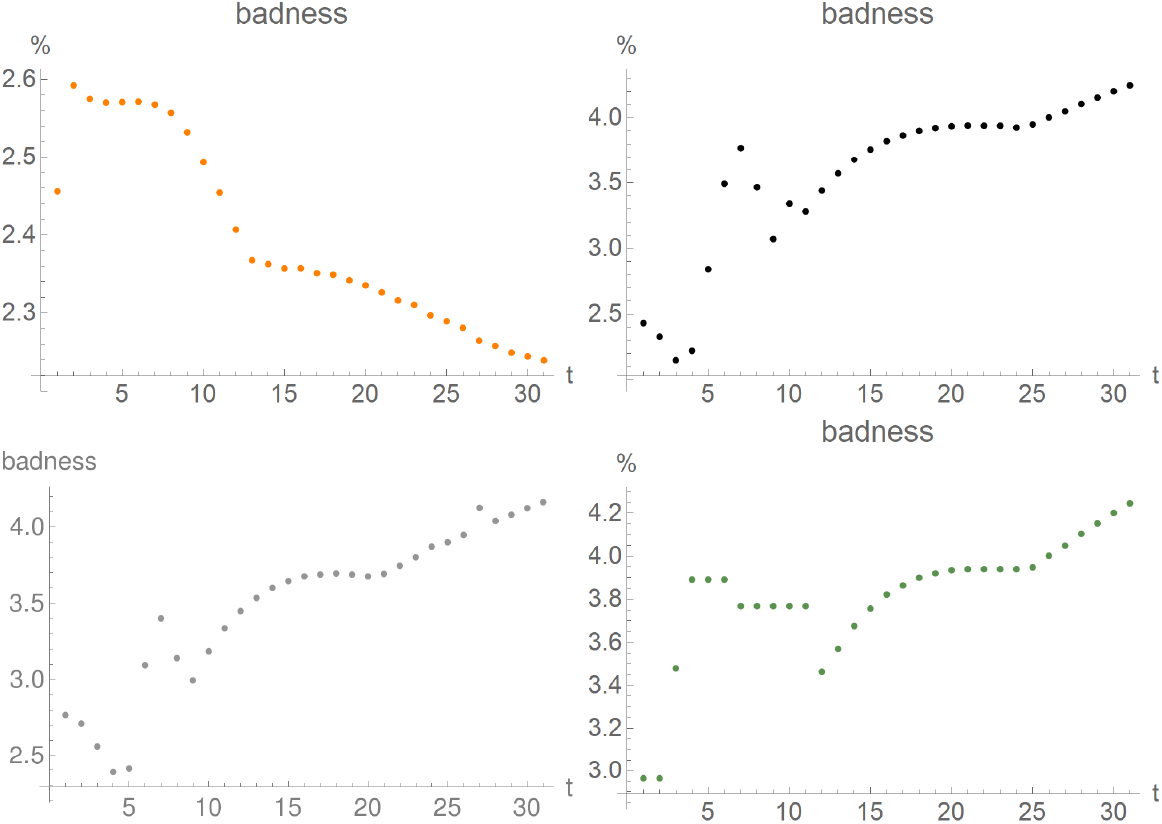
First “badness parameter” for fits for Austria generated in the period April 5 (day 1 on the plot) - May 5 (day 31 on the plot). Upper row: double logistic (left), logistic (right); lower row: Gauss-error (left), logistic fit with running constants (right).

**Figure 5.6:**
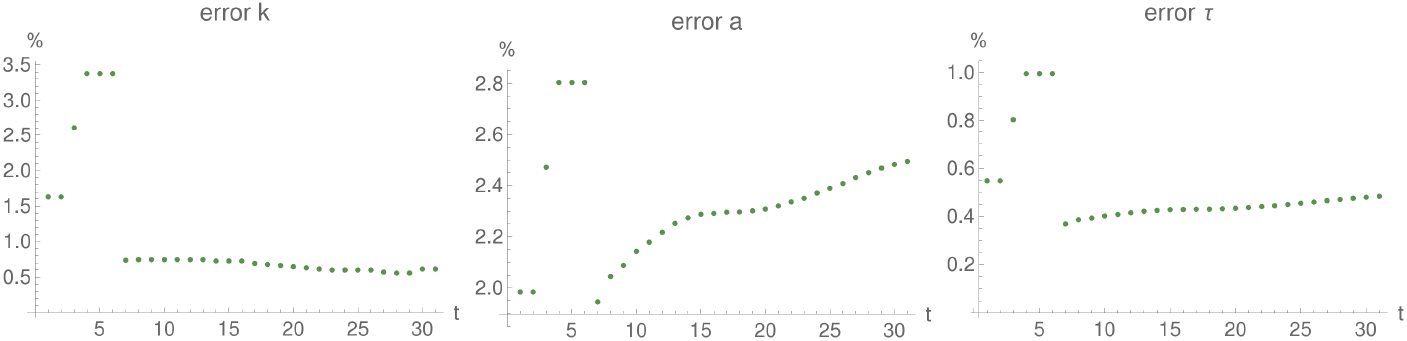
Maximal relative error of fit parameters for the logistic fit with running parameters for Austria, in the period April 5 (day 1 on the plot) - May 5 (day 31 on the plot).

## 6 Predictability of the algorithm

In order to obtain a global overview, we tested our algorithm on the time series available on the JHU server as follows. Recall, first, that the US time series, other than the collective one, are kept on the JHU server as a separate data set. We decided arbitrarily not to consider the US data set.

Next, from the remaining time series, we chose to consider only those where at least 2000 confirmed cases have been observed on May 5, 2020. It turns out that there are 72 such time series.

Now, there arose an issue related to the fact that Australia, Canada and China do not have global time series on the JHU server for the whole country, but separate ones for each sub-state or province. We decided, again arbitrarily, to exclude these time series from the analysis. Excluding China does not lead to any loss of information for the purposes of the analysis here in any case, since there the number of confirmed cases was essentially constant during the period under consideration.

We calculated all predictions for these time series. Their number turns out to be 32 055. From these predictions we arbitrarily decided to consider only these 16307 predictions with all badness parameters less than 10%. For these predictions we calculated, for each country, both the average and the maximal error for 1-day predictions (1197 predictions), 7-days predictions (802 predictions), and 14-days predictions (563 predictions).

Histograms showing the number of all predictions as above, within 1%-error bins, for all one-day, seven-days, and two-weeks predictions can be found in Figure 6.1.

**Figure 6.1:**
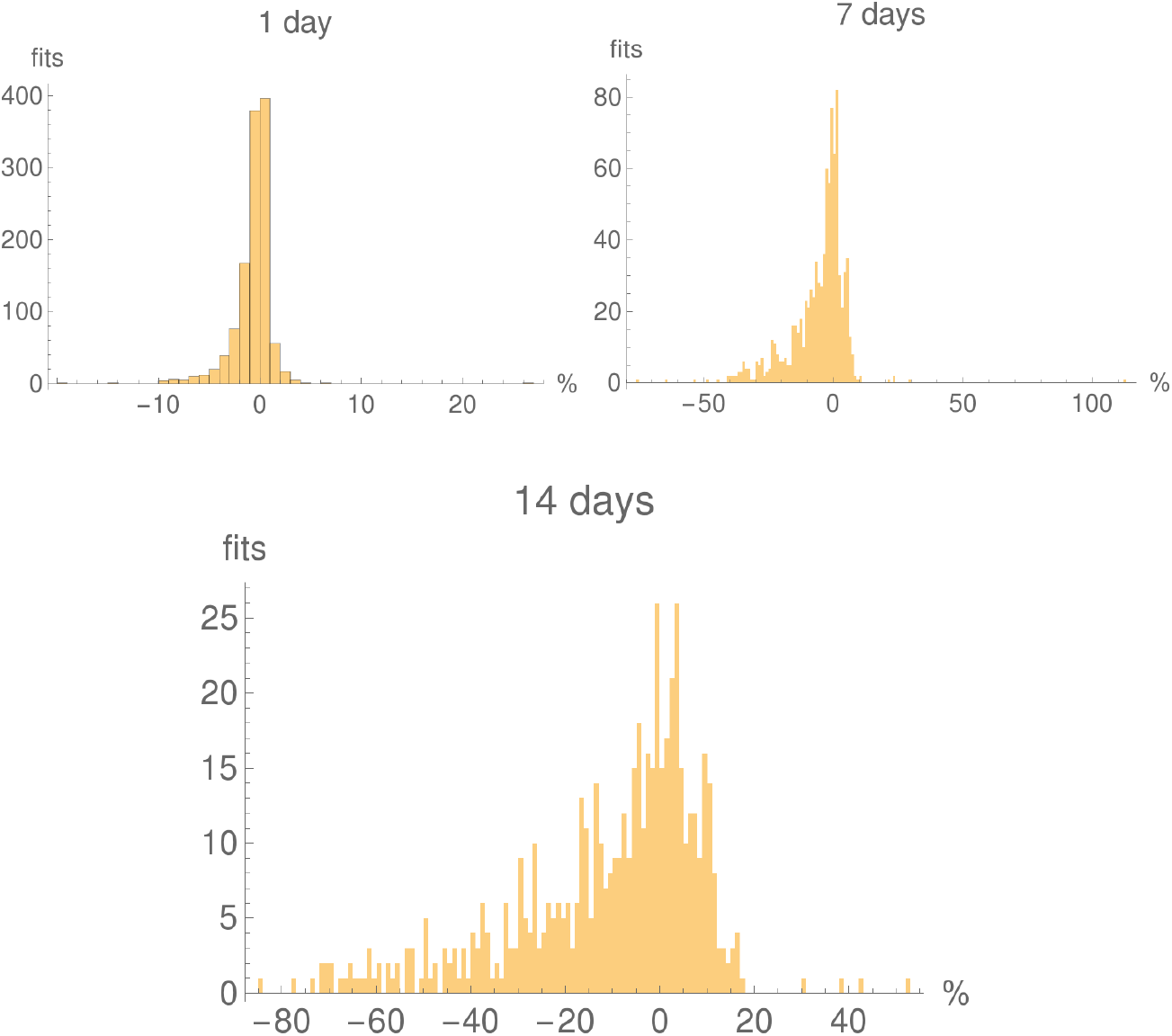
Histograms of the number of 1-day, 7-days and 2-weeks predictions with 1% error bins for selected countries, with tails cut-off. Note that all histograms have different vertical and horizontal scales.

**Figure 6.2:**
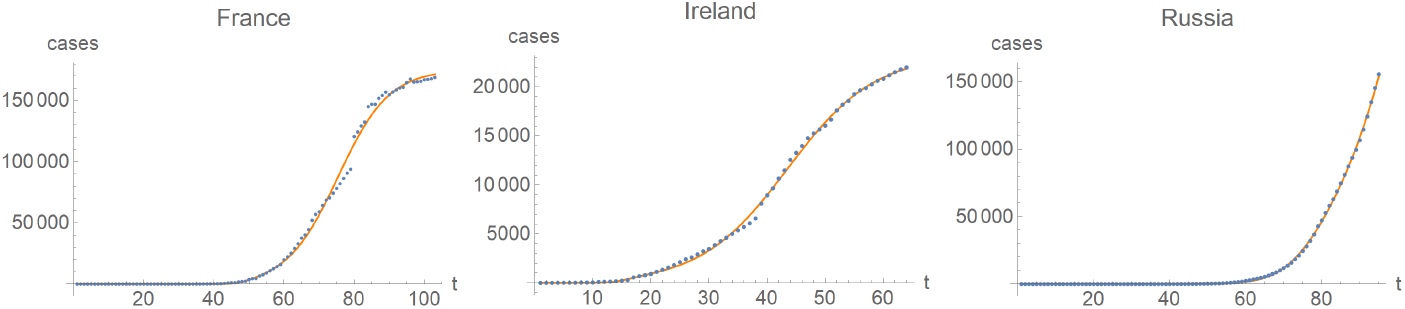
Double-logistic fits for France, Ireland and Russia on May 5, 2020.

We ordered our one-day, seven-days, and two-weeks predictions according to the average error of the prediction. The resulting lists are presented in Tables 6.1-6.3. (Some countries drop out from one list to the next when no fit meets the threshold for the longer period. Note that Cameroon and Singapore barely make it to Table 6.1, only one prediction meets all 10% thresholds there.) We emphasise that the ordering is only done to illustrate the effectiveness of our method, no other interpretations are implied.

**Table 6.1:** The average and maximal errors of one-day predictions, in percents, in the period April 5 - May 4 for selected countries. Only predictions with badness parameters smaller than 10% are kept. The ridiculously wrong one-day prediction for France and Ireland arise at the jumps in the time series, see Figure 6.2. The bad predictions for Russia correlate with fits with high badness parameters (around 8.8%) which we believe to be related to the fact that Russia is still in a relatively early phase of the epidemics.

|  | Country | av. error<br>[max. error] |  | Country | av. error<br>[max. error] |  | Country | av. error<br>[max. error] |
| --- | --- | --- | --- | --- | --- | --- | --- | --- |
| 1 | Korea, South | 0.185 [0.335] | 23 | Finland | 0.8 [3.23] | 45 | Chile | 1.56 [5.53] |
| 2 | Germany | 0.259 [1.28] | 24 | Poland | 0.807 [3.11] | 46 | South Africa | 1.71 [3.09] |
| 3 | Italy | 0.259 [0.984] | 25 | Belgium | 0.819 [3.85] | 47 | Mexico | 1.78 [3.99] |
| 4 | Iran | 0.291 [0.602] | 26 | Israel | 0.842 [2.46] | 48 | Iraq | 1.86 [4.12] |
| 5 | Austria | 0.308 [1.24] | 27 | Spain | 0.861 [6.51] | 49 | Dominican Rep. | 2.03 [6.6] |
| 6 | Switzerland | 0.343 [1.49] | 28 | Romania | 0.861 [3.56] | 50 | France | 2.04 [19.6] |
| 7 | Norway | 0.362 [1.41] | 29 | Serbia | 0.952 [2.1] | 51 | Armenia | 2.21 [4.4] |
| 8 | Thailand | 0.399 [1.77] | 30 | Portugal | 0.96 [6.33] | 52 | Morocco | 2.25 [7.33] |
| 9 | US | 0.442 [1.26] | 31 | Greece | 0.984 [6.16] | 53 | Japan | 2.44 [3.94] |
| 10 | Belarus | 0.468 [0.783] | 32 | Sweden | 1.11 [2.74] | 54 | Colombia | 2.47 [7.67] |
| 11 | Denmark | 0.52 [1.53] | 33 | Argentina | 1.12 [5.79] | 55 | Indonesia | 2.66 [9.05] |
| 12 | United Kingdom | 0.568 [2.01] | 34 | Turkey | 1.16 [4.72] | 56 | Afghanistan | 3.56 [9.04] |
| 13 | Malaysia | 0.579 [1.44] | 35 | Moldova | 1.26 [3.22] | 57 | Brazil | 3.7 [8.52] |
| 14 | United Arab Emir. | 0.597 [2.41] | 36 | Uzbekistan | 1.28 [2.79] | 58 | Kazakhstan | 3.72 [5.47] |
| 15 | Ukraine | 0.61 [1.1] | 37 | Algeria | 1.29 [3.89] | 59 | Bahrain | 3.81 [8.27] |
| 16 | Luxembourg | 0.618 [2.57] | 38 | Azerbaijan | 1.36 [5.22] | 60 | Saudi Arabia | 4. [6.54] |
| 17 | Netherlands | 0.648 [2.1] | 39 | Egypt | 1.46 [3.11] | 61 | Nigeria | 4.1 [8.11] |
| 18 | Oman | 0.658 [1.35] | 40 | India | 1.48 [5.13] | 62 | Peru | 4.31 [9.92] |
| 19 | Czechia | 0.682 [2.11] | 41 | Panama | 1.52 [5.34] | 63 | Pakistan | 5.05 [8.02] |
| 20 | Croatia | 0.701 [3.14] | 42 | Ireland | 1.55 [14.] | 64 | Russia | 5.58 [26.7] |
| 21 | Hungary | 0.744 [2.34] | 43 | Bangladesh | 1.55 [1.96] | 65 | Cameroon | 7.16 [7.16] |
| 22 | Philippines | 0.793 [1.64] | 44 | Singapore | 1.56 [1.56] |  |  |  |

**Table 6.2:** One-week predictions; the remaining details as in Table 6.1.

|  | Country | av. error<br>[max. error] |  | Country | av. error<br>[max. error] |  | Country | av. error<br>[max. error] |
| --- | --- | --- | --- | --- | --- | --- | --- | --- |
| 1 | Ukraine | 0.734 [1.39] | 22 | Spain | 4.03 [10.1] | 43 | Morocco | 16.1 [23.5] |
| 2 | Germany | 0.84 [2.56] | 23 | Netherlands | 4.31 [8.14] | 44 | Japan | 16.5 [18.4] |
| 3 | Italy | 1.18 [2.26] | 24 | Croatia | 4.6 [10.4] | 45 | Uzbekistan | 16.8 [32.2] |
| 4 | Korea, South | 1.7 [2.5] | 25 | Poland | 4.67 [21.8] | 46 | Colombia | 16.9 [29.9] |
| 5 | Thailand | 1.91 [7.41] | 26 | Denmark | 5.21 [10.4] | 47 | Armenia | 17.1 [23.4] |
| 6 | Norway | 1.92 [5.48] | 27 | Greece | 5.48 [29.5] | 48 | Mexico | 18.6 [18.6] |
| 7 | Austria | 1.94 [6.12] | 28 | United Arab Emir. | 5.62 [7.64] | 49 | Iraq | 18.8 [29.] |
| 8 | Romania | 1.94 [13.8] | 29 | Sweden | 5.66 [16.6] | 50 | France | 19.3 [34.7] |
| 9 | Iran | 2.33 [3.41] | 30 | Panama | 6.57 [17.8] | 51 | Pakistan | 20.7 [22.1] |
| 10 | United Kingdom | 2.4 [4.72] | 31 | Moldova | 6.84 [12.7] | 52 | Brazil | 20.8 [33.5] |
| 11 | Switzerland | 2.4 [7.2] | 32 | Serbia | 7.32 [12.] | 53 | Indonesia | 21.6 [34.6] |
| 12 | US | 2.64 [5.32] | 33 | Argentina | 8.16 [13.8] | 54 | Cameroon | 23.1 [23.1] |
| 13 | Luxembourg | 2.64 [7.86] | 34 | Chile | 8.37 [21.1] | 55 | Afghanistan | 24.5 [39.2] |
| 14 | Portugal | 3.19 [10.7] | 35 | Turkey | 9.23 [27.2] | 56 | Peru | 25. [29.3] |
| 15 | Israel | 3.38 [10.6] | 36 | India | 9.56 [15.2] | 57 | Kazakhstan | 33.8 [33.9] |
| 16 | Czechia | 3.41 [7.82] | 37 | Egypt | 10.5 [20.4] | 58 | Nigeria | 34.9 [48.4] |
| 17 | Malaysia | 3.65 [7.23] | 38 | Algeria | 11.4 [20.9] | 59 | Saudi Arabia | 35.4 [40.] |
| 18 | Philippines | 3.66 [6.7] | 39 | Dominican Rep. | 11.9 [29.6] | 60 | Bahrain | 36.7 [39.6] |
| 19 | Hungary | 3.8 [9.45] | 40 | Ireland | 12.5 [31.4] | 61 | Russia | 45.3 [112.] |
| 20 | Finland | 3.86 [12.5] | 41 | Azerbaijan | 14. [40.3] |  |  |  |
| 21 | Belgium | 4.02 [9.7] | 42 | South Africa | 15.8 [17.] |  |  |  |

**Table 6.3:** Two-week predictions; the remaining details as in Table 6.1.

|  | Country | av. error<br>[max. error] |  | Country | av. error<br>[max. error] |  | Country | av. error<br>[max. error] |
| --- | --- | --- | --- | --- | --- | --- | --- | --- |
| 1 | Italy | 1.7 [3.29] | 20 | Malaysia | 8.82 [13.4] | 39 | Armenia | 31.5 [37.4] |
| 2 | Germany | 1.74 [2.86] | 21 | Finland | 10.4 [23.4] | 40 | Dominican Rep. | 32. [65.6] |
| 3 | Portugal | 3.91 [16.1] | 22 | Greece | 10.8 [52.8] | 41 | Mexico | 32.1 [32.1] |
| 4 | Austria | 4.3 [7.83] | 23 | Moldova | 11.7 [16.3] | 42 | France | 33.3 [49.7] |
| 5 | Korea, South | 4.38 [5.62] | 24 | Poland | 11.8 [31.] | 43 | Brazil | 36.4 [36.4] |
| 6 | Thailand | 4.45 [16.6] | 25 | Serbia | 12.9 [17.4] | 44 | Pakistan | 37.6 [38.8] |
| 7 | United Kingdom | 4.58 [10.] | 26 | Philippines | 13.9 [19.2] | 45 | Afghanistan | 39.8 [47.3] |
| 8 | Norway | 4.62 [9.38] | 27 | Hungary | 14.8 [17.3] | 46 | Ireland | 40.6 [44.5] |
| 9 | Luxembourg | 4.72 [10.] | 28 | Panama | 15. [27.] | 47 | Indonesia | 41.7 [49.2] |
| 10 | Spain | 5.22 [11.1] | 29 | Egypt | 15.6 [19.2] | 48 | South Africa | 41.8 [53.9] |
| 11 | Romania | 5.29 [26.8] | 30 | Chile | 17.8 [28.8] | 49 | Azerbaijan | 43.4 [84.1] |
| 12 | Israel | 6.28 [14.6] | 31 | Denmark | 19.1 [22.] | 50 | Bahrain | 44.4 [49.2] |
| 13 | Iran | 6.56 [7.94] | 32 | United Arab Emir. | 19.2 [19.2] | 51 | Cameroon | 47.2 [47.2] |
| 14 | Belgium | 6.58 [17.1] | 33 | Argentina | 19.4 [27.1] | 52 | Iraq | 48.8 [62.2] |
| 15 | US | 6.78 [13.7] | 34 | Sweden | 20.1 [37.1] | 53 | Russia | 52.3 [55.1] |
| 16 | Croatia | 6.93 [10.4] | 35 | Turkey | 23.5 [52.2] | 54 | Kazakhstan | 52.7 [52.8] |
| 17 | Czechia | 7.67 [15.1] | 36 | Algeria | 27.3 [38.7] | 55 | Colombia | 53.3 [53.3] |
| 18 | Switzerland | 7.7 [15.2] | 37 | India | 27.9 [29.8] | 56 | Nigeria | 65.4 [73.6] |
| 19 | Netherlands | 8.01 [13.5] | 38 | Morocco | 28.8 [32.8] | 57 | Saudi Arabia | 69.4 [71.5] |

It is legitimate to raise the question, how otherwise reasonable fits can lead to very poor predictions, be it for one or more days. Such cases arise when the rates of increase of the *k* parameter exhibit significant variations. This can be due to a wobbly epidemic curve which results from a lack of systematic or consistent reporting of new cases, or a change in testing practices. Barring that, this can also result from a real decrease of the dynamics the epidemics, e.g. from the introduction of stricter confinement measures. In any case, our algorithm is only tailored to periods where the rates of change of the fitting parameters do not vary much in time.

For comparison we also present in Table 6.4 the analysis of the 20888 predictions with all badness parameters less than 20% (where Qatar dropped out from the list of 69 countries considered), and the 7840 predictions with badness parameters less than 5% in Table 6.5.

**Table 6.4:** The average error of one-day predictions, in percents, with all badness parameters smaller than 20%. The remaining details as in Table 6.1.

|  | Country | av. error<br>[max. error] |  | Country | av. error<br>[max. error] |  | Country | av. error<br>[max. error] |
| --- | --- | --- | --- | --- | --- | --- | --- | --- |
| 1 | Korea, South | 0.185 [0.335] | 24 | Serbia | 0.952 [2.1] | 47 | Morocco | 2.04 [7.33] |
| 2 | Germany | 0.259 [1.28] | 25 | Portugal | 0.96 [6.33] | 48 | France | 2.22 [19.6] |
| 3 | Italy | 0.259 [0.984] | 26 | Greece | 0.984 [6.16] | 49 | Armenia | 2.42 [4.67] |
| 4 | Austria | 0.308 [1.24] | 27 | Argentina | 1.12 [5.79] | 50 | Japan | 2.44 [3.94] |
| 5 | Switzerland | 0.343 [1.49] | 28 | Sweden | 1.12 [2.74] | 51 | Colombia | 2.54 [7.67] |
| 6 | Norway | 0.362 [1.41] | 29 | Turkey | 1.16 [4.72] | 52 | Indonesia | 2.57 [9.05] |
| 7 | Thailand | 0.399 [1.77] | 30 | Finland | 1.16 [7.72] | 53 | Kazakhstan | 2.78 [12.] |
| 8 | US | 0.442 [1.26] | 31 | Moldova | 1.24 [3.22] | 54 | Mexico | 3.31 [9.74] |
| 9 | Denmark | 0.52 [1.53] | 32 | Belarus | 1.24 [2.57] | 55 | South Africa | 3.32 [7.82] |
| 10 | Malaysia | 0.579 [1.44] | 33 | Algeria | 1.29 [3.89] | 56 | Afghanistan | 3.46 [9.04] |
| 11 | Luxembourg | 0.618 [2.57] | 34 | Ukraine | 1.35 [8.72] | 57 | Cameroon | 3.72 [10.8] |
| 12 | Netherlands | 0.648 [2.1] | 35 | India | 1.48 [5.13] | 58 | Brazil | 3.88 [8.52] |
| 13 | Czechia | 0.682 [2.11] | 36 | Uzbekistan | 1.5 [4.37] | 59 | Oman | 3.89 [14.3] |
| 14 | Croatia | 0.701 [3.14] | 37 | Azerbaijan | 1.5 [5.22] | 60 | Peru | 3.91 [9.92] |
| 15 | United Kingdom | 0.715 [4.21] | 38 | Panama | 1.52 [5.34] | 61 | Pakistan | 4.17 [14.2] |
| 16 | United Arab Emir. | 0.755 [2.41] | 39 | Chile | 1.53 [5.53] | 62 | Kuwait | 4.39 [6.] |
| 17 | Philippines | 0.793 [1.64] | 40 | Bangladesh | 1.55 [1.96] | 63 | Bahrain | 4.71 [14.3] |
| 18 | Belgium | 0.819 [3.85] | 41 | Singapore | 1.56 [1.56] | 64 | Saudi Arabia | 4.72 [7.04] |
| 19 | Iran | 0.838 [9.25] | 42 | Iraq | 1.78 [4.12] | 65 | Nigeria | 5.15 [13.1] |
| 20 | Israel | 0.842 [2.46] | 43 | Egypt | 1.8 [7.56] | 66 | Russia | 5.58 [26.7] |
| 21 | Spain | 0.861 [6.51] | 44 | Hungary | 1.83 [14.5] | 67 | Ghana | 7.07 [23.7] |
| 22 | Romania | 0.861 [3.56] | 45 | Dominican Rep. | 1.98 [6.6] | 68 | Ecuador | 10.6 [31.8] |
| 23 | Poland | 0.949 [3.61] | 46 | Ireland | 1.98 [14.] |  |  |  |

**Table 6.5:** The average error of one-day predictions, in percents, with all badness parameters smaller than 5%. The remaining details as in Table 6.1.

|  | Country | av. error<br>[max. error] |  | Country | av. error<br>[max. error] |  | Country | av. error<br>[max. error] |
| --- | --- | --- | --- | --- | --- | --- | --- | --- |
| 1 | Germany | 0.259 [1.28] | 18 | Ukraine | 0.612 [1.09] | 35 | Dominican Rep. | 1.44 [2.29] |
| 2 | Turkey | 0.26 [0.655] | 19 | Luxembourg | 0.634 [1.16] | 36 | Egypt | 1.45 [2.1] |
| 3 | Italy | 0.285 [0.984] | 20 | Netherlands | 0.648 [2.1] | 37 | Iraq | 1.45 [2.04] |
| 4 | Iran | 0.291 [0.602] | 21 | Oman | 0.658 [1.35] | 38 | Bangladesh | 1.55 [1.96] |
| 5 | Austria | 0.308 [1.24] | 22 | Croatia | 0.668 [3.14] | 39 | Singapore | 1.56 [1.56] |
| 6 | Norway | 0.332 [1.41] | 23 | Belgium | 0.713 [2.57] | 40 | Panama | 1.8 [3.91] |
| 7 | United Arab Emir. | 0.338 [0.566] | 24 | Romania | 0.753 [3.39] | 41 | Moldova | 1.81 [3.22] |
| 8 | Switzerland | 0.343 [1.49] | 25 | Azerbaijan | 0.774 [1.42] | 42 | Morocco | 1.97 [1.97] |
| 9 | Thailand | 0.399 [1.77] | 26 | Indonesia | 0.784 [1.4] | 43 | Russia | 2. [3.19] |
| 10 | Poland | 0.402 [1.15] | 27 | Spain | 0.832 [2.19] | 44 | Algeria | 2.03 [2.03] |
| 11 | United Kingdom | 0.478 [1.27] | 28 | Sweden | 0.859 [1.92] | 45 | Argentina | 2.05 [5.79] |
| 12 | US | 0.484 [1.26] | 29 | Greece | 0.895 [6.16] | 46 | Armenia | 2.2 [2.24] |
| 13 | Czechia | 0.499 [2.11] | 30 | Serbia | 0.963 [2.1] | 47 | Nigeria | 2.48 [2.48] |
| 14 | Hungary | 0.513 [0.979] | 31 | Chile | 0.968 [4.26] | 48 | Mexico | 3.99 [3.99] |
| 15 | Denmark | 0.522 [1.53] | 32 | Portugal | 1.05 [6.33] | 49 | Peru | 9.92 [9.92] |
| 16 | Finland | 0.555 [1.3] | 33 | India | 1.25 [2.49] | 50 | Ireland | 14. [14.] |
| 17 | Malaysia | 0.612 [1.44] | 34 | Israel | 1.31 [1.34] | 51 | France | 19.6 [19.6] |

The results of the above analysis restricted to EU countries can be found in Figure 6.3 and in Tables 6.6-6.10.

**Figure 6.3:**
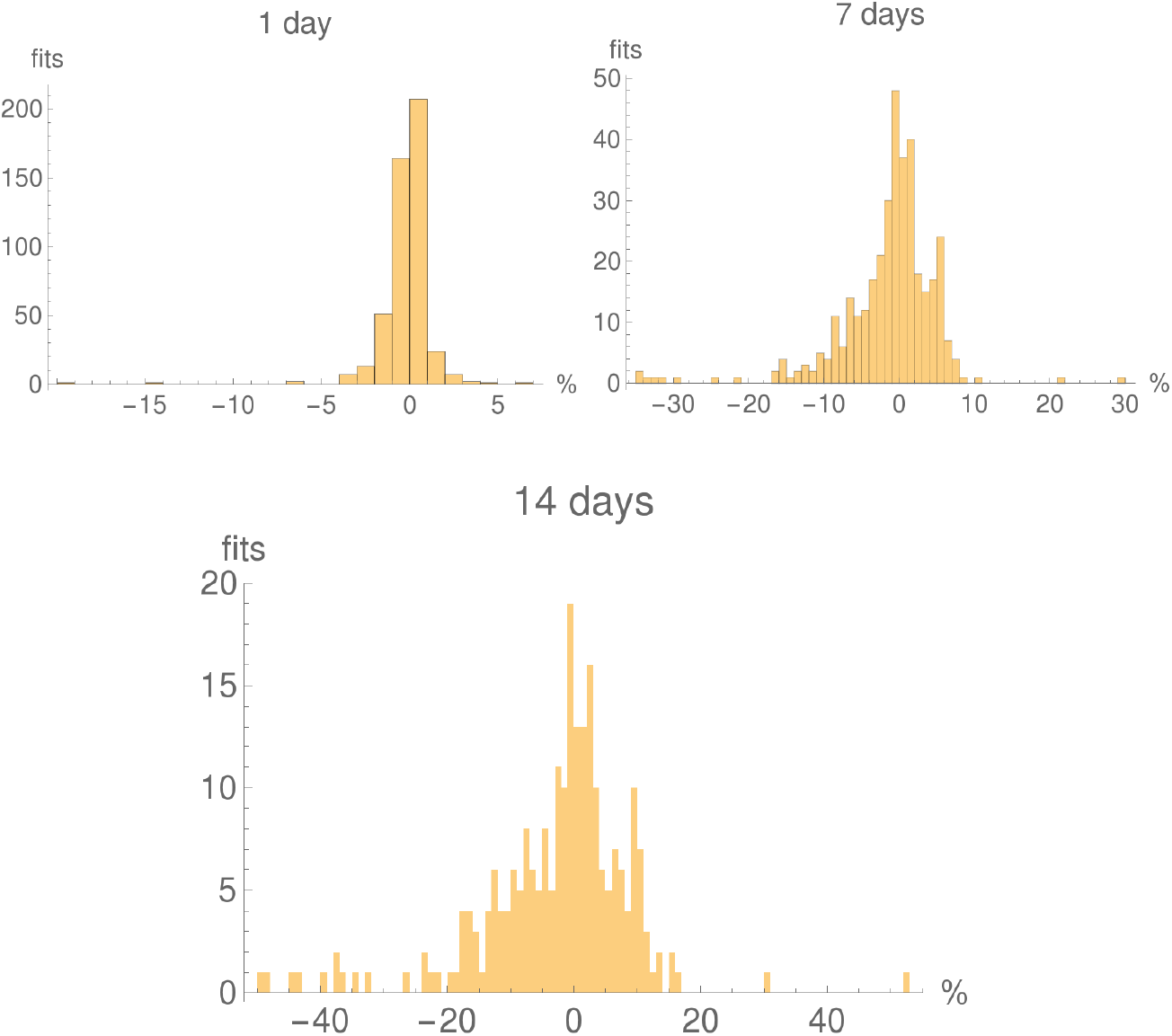
Histograms of the number of 1-day, 7-days and 2-weeks predictions with 1% error bins for those EU countries which had more than 2000 confirmed cases on May 5. The tails of the histograms are cut-off. All histograms have different vertical and horizontal scales.

**Table 6.6:** The average and maximal errors of one-day predictions, in percents, in the period April 5 - May 4 for EU countries. All badness parameters smaller than 10%. As noted previously, the ridiculously wrong one-day prediction for France arises at the day of the jump in the French time series.

|  | Country | av. error<br>[max. error] |  | Country | av. error<br>[max. error] |  | Country | av. error<br>[max. error] |
| --- | --- | --- | --- | --- | --- | --- | --- | --- |
| 1 | Germany | 0.259 [1.28] | 8 | Croatia | 0.701 [3.14] | 15 | Portugal | 0.96 [6.33] |
| 2 | Italy | 0.259 [0.984] | 9 | Hungary | 0.744 [2.34] | 16 | Greece | 0.984 [6.16] |
| 3 | Austria | 0.308 [1.24] | 10 | Finland | 0.8 [3.23] | 17 | Sweden | 1.11 [2.74] |
| 4 | Denmark | 0.52 [1.53] | 11 | Poland | 0.807 [3.11] | 18 | Ireland | 1.55 [14.] |
| 5 | Luxembourg | 0.618 [2.57] | 12 | Belgium | 0.819 [3.85] | 19 | France | 2.04 [19.6] |
| 6 | Netherlands | 0.648 [2.1] | 13 | Spain | 0.861 [6.51] |  |  |  |
| 7 | Czechia | 0.682 [2.11] | 14 | Romania | 0.861 [3.56] |  |  |  |

**Table 6.7:** One-weeks predictions; the remaining details as in Table 6.6.

|  | Country | av. error<br>[max. error] |  | Country | av. error<br>[max. error] |  | Country | av. error<br>[max. error] |
| --- | --- | --- | --- | --- | --- | --- | --- | --- |
| 1 | Germany | 0.84 [2.56] | 8 | Hungary | 3.8 [9.45] | 15 | Denmark | 5.21 [10.4] |
| 2 | Italy | 1.18 [2.26] | 9 | Finland | 3.86 [12.5] | 16 | Greece | 5.48 [29.5] |
| 3 | Austria | 1.94 [6.12] | 10 | Belgium | 4.02 [9.7] | 17 | Sweden | 5.66 [16.6] |
| 4 | Romania | 1.94 [13.8] | 11 | Spain | 4.03 [10.1] | 18 | Ireland | 12.5 [31.4] |
| 5 | Luxembourg | 2.64 [7.86] | 12 | Netherlands | 4.31 [8.14] | 19 | France | 19.3 [34.7] |
| 6 | Portugal | 3.19 [10.7] | 13 | Croatia | 4.6 [10.4] |  |  |  |
| 7 | Czechia | 3.41 [7.82] | 14 | Poland | 4.67 [21.8] |  |  |  |

**Table 6.8:** Two-weeks predictions; the remaining details as in Table 6.6.

|  | Country | av. error<br>[max. error] |  | Country | av. error<br>[max. error] |  | Country | av. error<br>[max. error] |
| --- | --- | --- | --- | --- | --- | --- | --- | --- |
| 1 | Italy | 1.7 [3.29] | 8 | Belgium | 6.58 [17.1] | 15 | Hungary | 14.8 [17.3] |
| 2 | Germany | 1.74 [2.86] | 9 | Croatia | 6.93 [10.4] | 16 | Denmark | 19.1 [22.] |
| 3 | Portugal | 3.91 [16.1] | 10 | Czechia | 7.67 [15.1] | 17 | Sweden | 20.1 [37.1] |
| 4 | Austria | 4.3 [7.83] | 11 | Netherlands | 8.01 [13.5] | 18 | France | 33.3 [49.7] |
| 5 | Luxembourg | 4.72 [10.] | 12 | Finland | 10.4 [23.4] | 19 | Ireland | 40.6 [44.5] |
| 6 | Spain | 5.22 [11.1] | 13 | Greece | 10.8 [52.8] |  |  |  |
| 7 | Romania | 5.29 [26.8] | 14 | Poland | 11.8 [31.] |  |  |  |

**Table 6.9:**
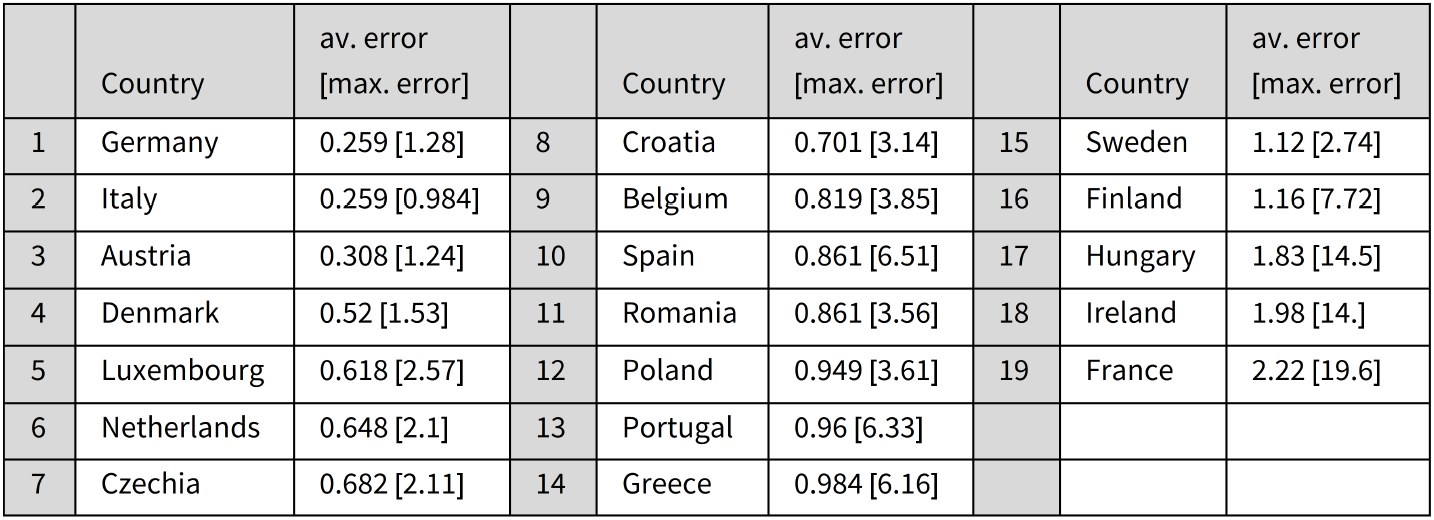
One-day fits with badness parameters smaller than 20%. The remaining details as in Table 6.6.

|  | Country | av. error<br>[max. error] |  | Country | av. error<br>[max. error] |  | Country | av. error<br>[max. error] |
| --- | --- | --- | --- | --- | --- | --- | --- | --- |
| 1 | Germany | 0.259 [1.28] | 8 | Croatia | 0.701 [3.14] | 15 | Sweden | 1.12 [2.74] |
| 2 | Italy | 0.259 [0.984] | 9 | Belgium | 0.819 [3.85] | 16 | Finland | 1.16 [7.72] |
| 3 | Austria | 0.308 [1.24] | 10 | Spain | 0.861 [6.51] | 17 | Hungary | 1.83 [14.5] |
| 4 | Denmark | 0.52 [1.53] | 11 | Romania | 0.861 [3.56] | 18 | Ireland | 1.98 [14.] |
| 5 | Luxembourg | 0.618 [2.57] | 12 | Poland | 0.949 [3.61] | 19 | France | 2.22 [19.6] |
| 6 | Netherlands | 0.648 [2.1] | 13 | Portugal | 0.96 [6.33] |  |  |  |
| 7 | Czechia | 0.682 [2.11] | 14 | Greece | 0.984 [6.16] |  |  |  |

**Table 6.10:** One-day fits with badness parameters smaller than 5%. The remaining details as in Table 6.6.

|  | Country | av. error<br>[max. error] |  | Country | av. error<br>[max. error] |  | Country | av. error<br>[max. error] |
| --- | --- | --- | --- | --- | --- | --- | --- | --- |
| 1 | Germany | 0.259 [1.28] | 8 | Finland | 0.555 [1.3] | 15 | Sweden | 0.859 [1.92] |
| 2 | Italy | 0.285 [0.984] | 9 | Luxembourg | 0.634 [1.16] | 16 | Greece | 0.895 [6.16] |
| 3 | Austria | 0.308 [1.24] | 10 | Netherlands | 0.648 [2.1] | 17 | Portugal | 1.05 [6.33] |
| 4 | Poland | 0.402 [1.15] | 11 | Croatia | 0.668 [3.14] | 18 | Ireland | 14. [14.] |
| 5 | Czechia | 0.499 [2.11] | 12 | Belgium | 0.713 [2.57] | 19 | France | 19.6 [19.6] |
| 6 | Hungary | 0.513 [0.979] | 13 | Romania | 0.753 [3.39] |  |  |  |
| 7 | Denmark | 0.522 [1.53] | 14 | Spain | 0.832 [2.19] |  |  |  |

**Figure A.1:**
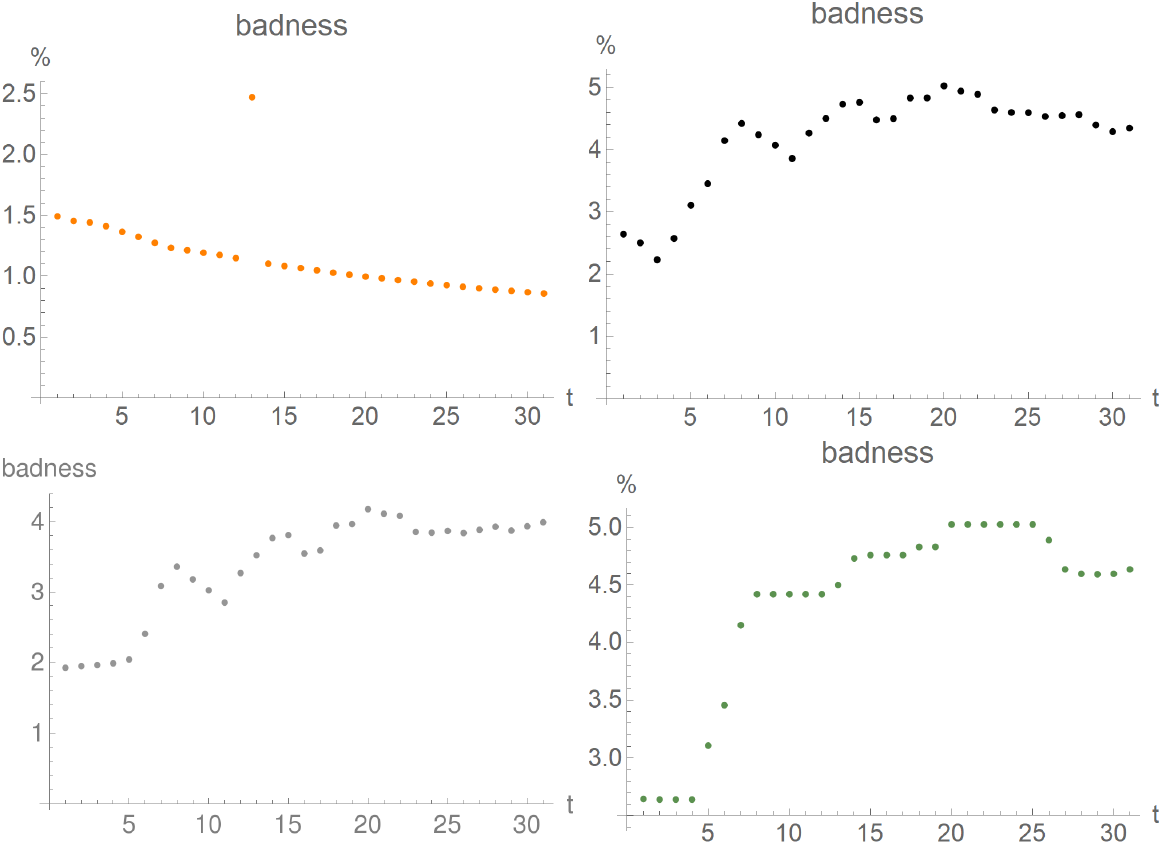
First “badness parameter” for fits for Italy generated in the period April 5 (day 1 on the plot) - May 5 (day 31 on the plot). Upper row: double logistic (left), logistic (right); lower row: Gauss-error (left), logistic with running constants (right).

## 7 Conclusions

We have shown that a phenomenological curve, the “double-logistic” five-parameter curve, can be used to describe the Covid-19 epidemics with reasonable accuracy at any given moment of time. We have analysed how the logistic, double-logistic, and Gauss-error models can be used for short-term predictions for the evolution of the current phase of the epidemics. We have presented an algorithm, based on the logistic curve, which is moderately successful in short-term predictions for a range of countries.

It is clear that neither the curves analysed, nor the algorithm, are suitable for long-term predictions of the epidemics.

We will be glad to provide the results of a similar analysis for other countries, or other fitting periods, upon request.

## Data Availability

All data underlying this research are publicly available at the John Hopkins University server. All further details of our analysis are available upon request.

## A Further selected data sets

### A.1 Italy

In this section we present the results of our analysis for Italy, in parallel to the presentation of Austria in Section 5.

The plots of the first badness parameter are found in Figure A.1, while the relative errors of the fits can be found in Figure A.2.

The fits with their standard errors and residuals are found in Figure A.3.

In Figure A.4 we show how the parameters of the fits, and their confidence intervals, change in time, for the double logistic, Gauss-error, and logistic curves. The predictability strings for Italy are plotted in Figure A.5.

**Figure A.2:**
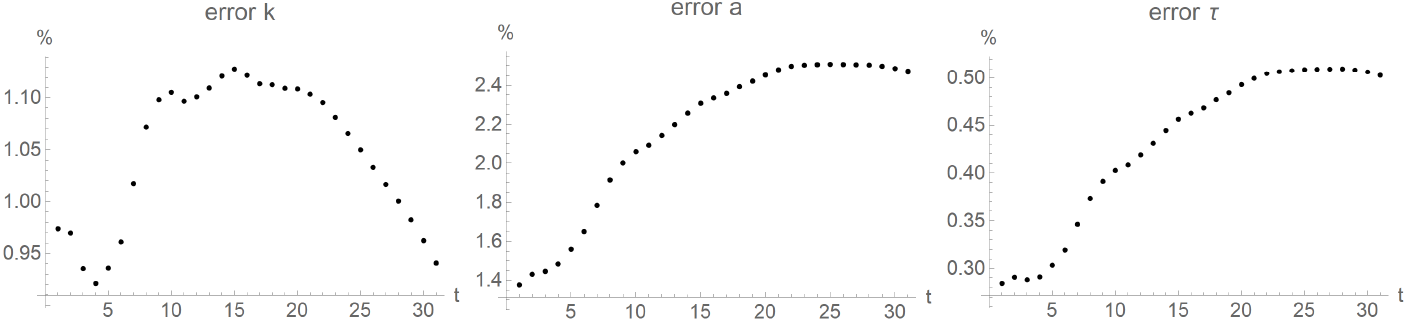
Relative error of logistic-fit parameters for Italy generated in the period April 5 (day 1 on the plot) - May 5 (day 31 on the plot).

In Figure A.6 we show the histograms of the errors of the predictions.

**Figure A.3:**
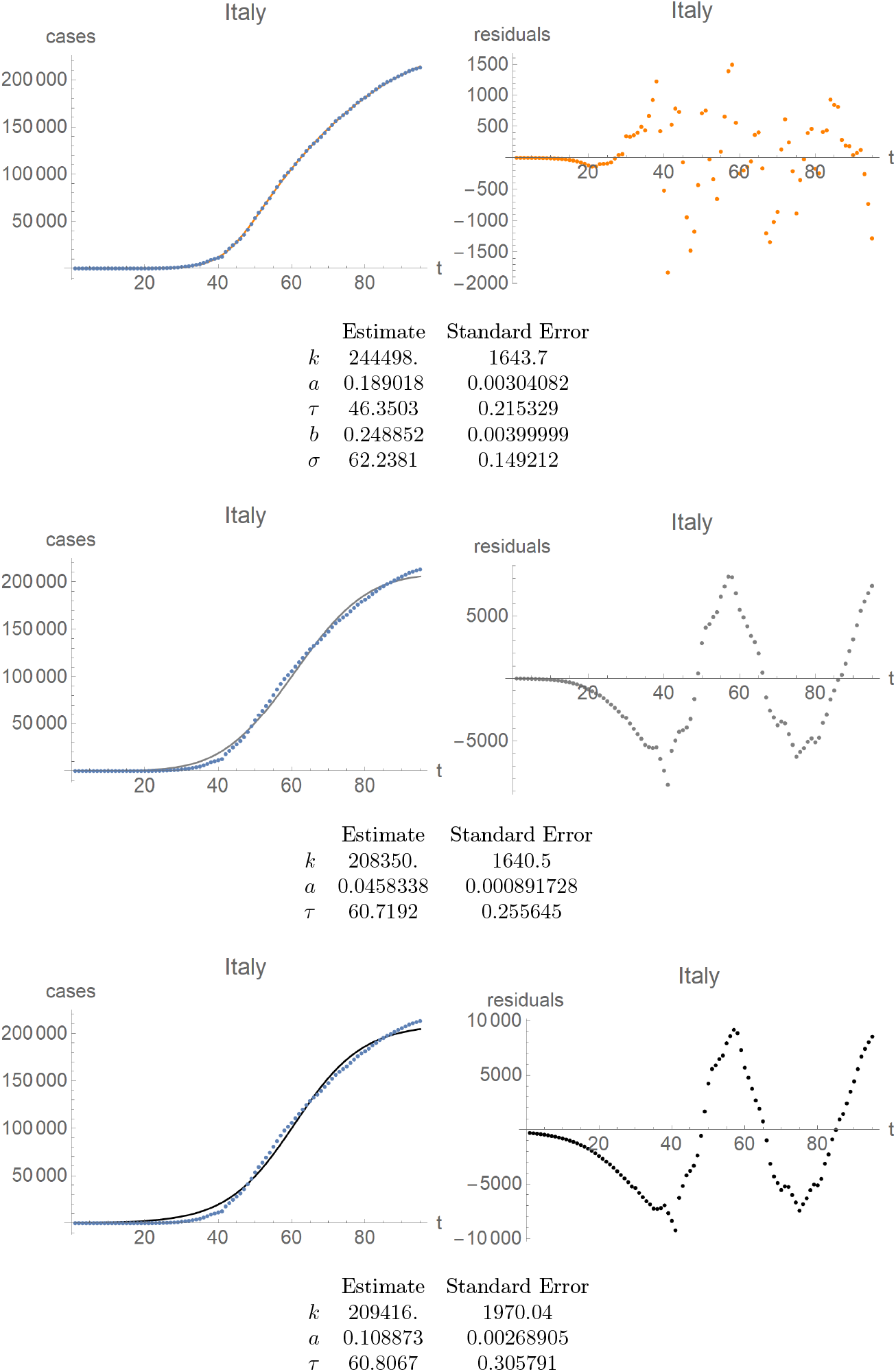
Italy. First row: fit of the double-logistic curve (left) and its residuals (right). Same for the Gauss-error curve in the second row, and the logistic curves in the third row. Note distinct scales for the residual plots as compared to the fitted curve, as well as a distinct scale for the residuals of the double-logistic curve compared to the other residuals.

**Figure A.4:**
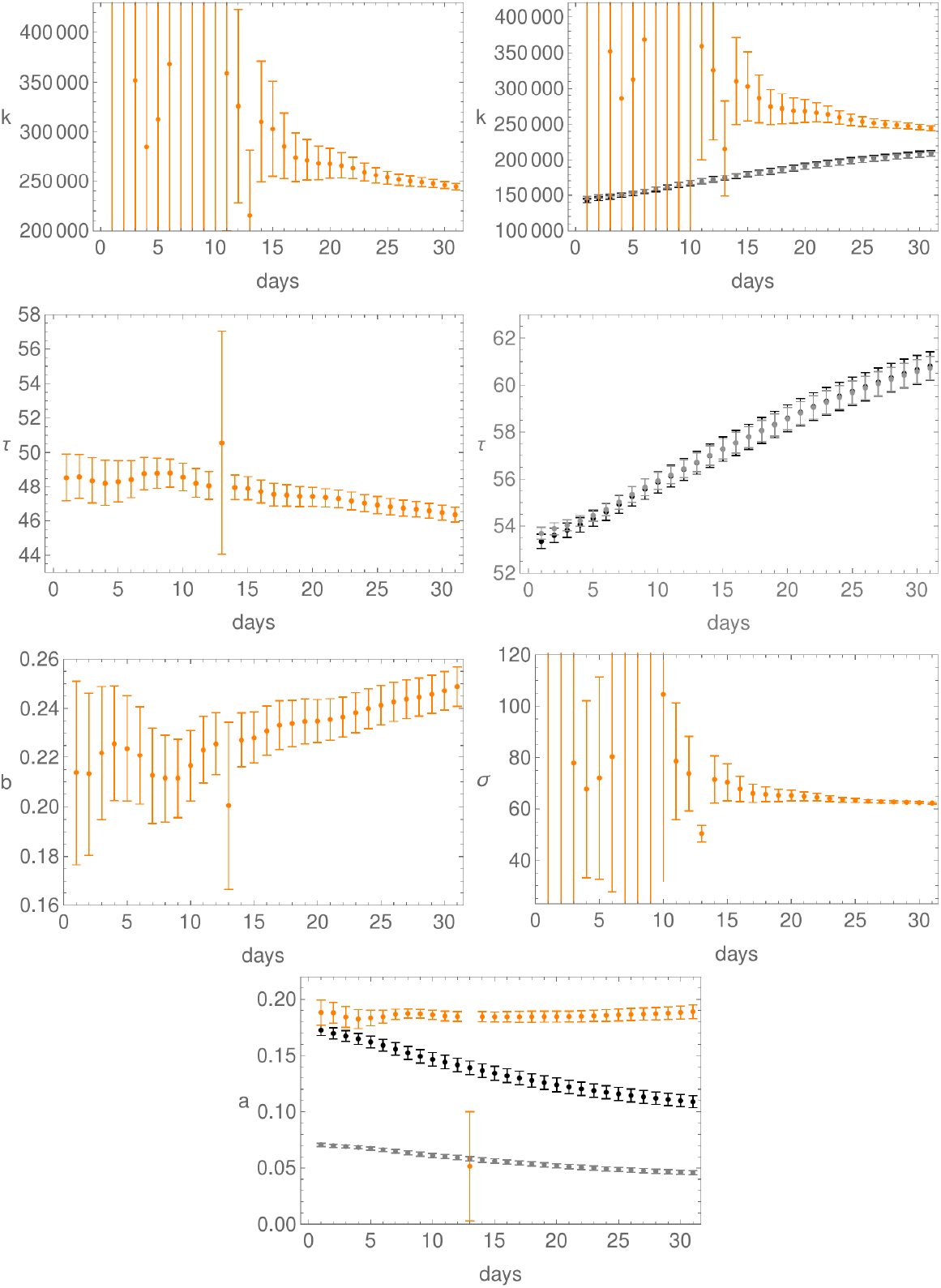
Evolution of the fit parameters in time, together with their confidence intervals, for Italy; day 1 is April 5. First row: parameter *k* for the double-logistic curve (left), and for all three curves (right). Here, as elsewhere, orange is for the double-logistic, black for the logistic, and grey for the Gauss-error function. Second row: the *τ*-parameter for the double-logistic curve (left) and for both the logistic and Gauss-error curves (right). Third row: plots of the parameters *b* (left) and *σ* (right) for the double-logistic curve. Last row: plot of the parameter *a* for all three curves.

**Figure A.5:**
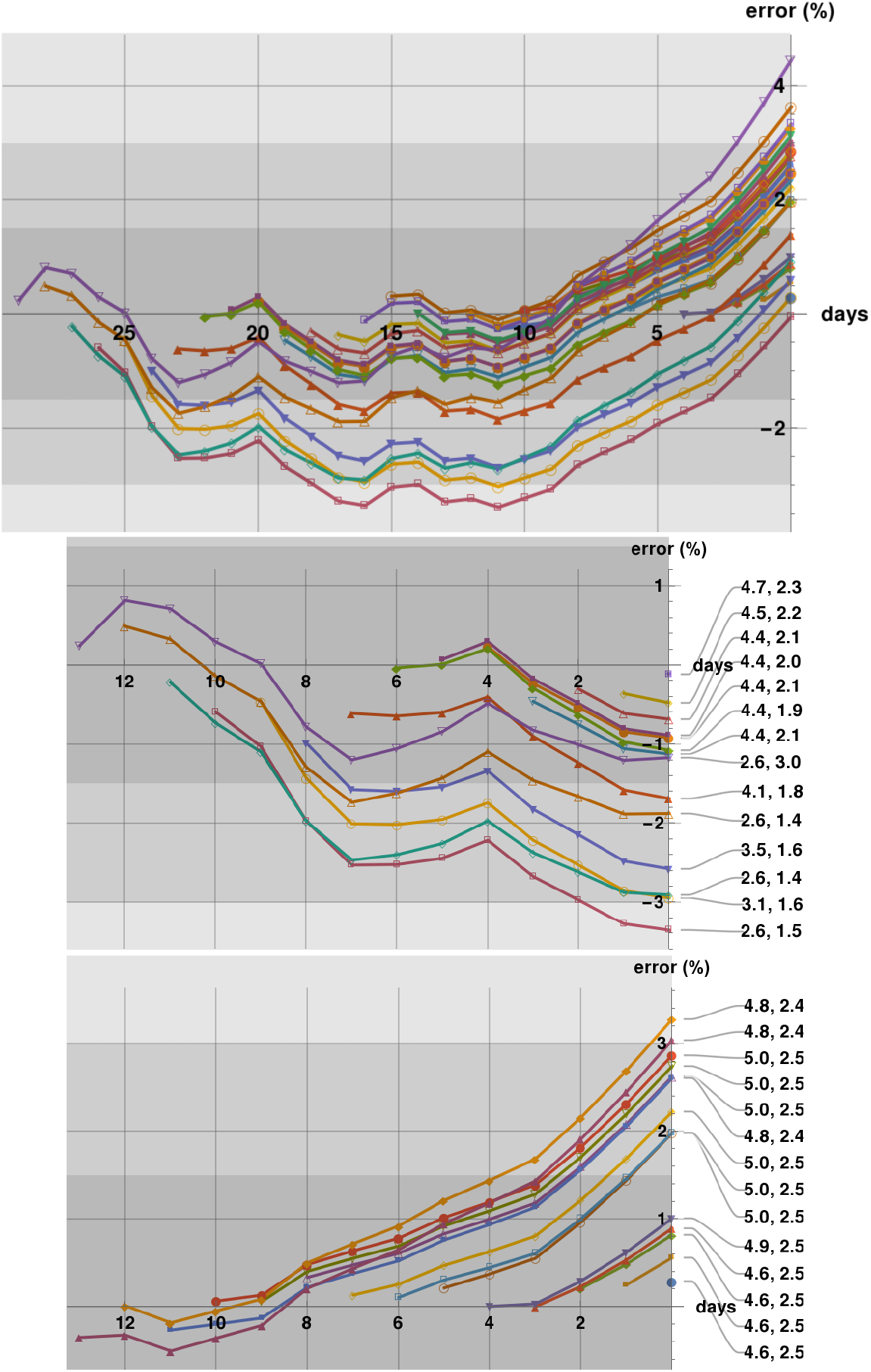
Predictability plots for Italy generated in the period April 5 - May 4. The first row shows all predictability strings with last prediction on May 5, which is the last day on the first and third rows. The shadings correspond to a 1.5%, 3%, and 6% error zones. The second row shows the predictability strings cut-off at April 20, with badness parameters indicated. The third row shows only these predictability strings which start within a two-weeks period ending on May 5.

**Figure A.6:**
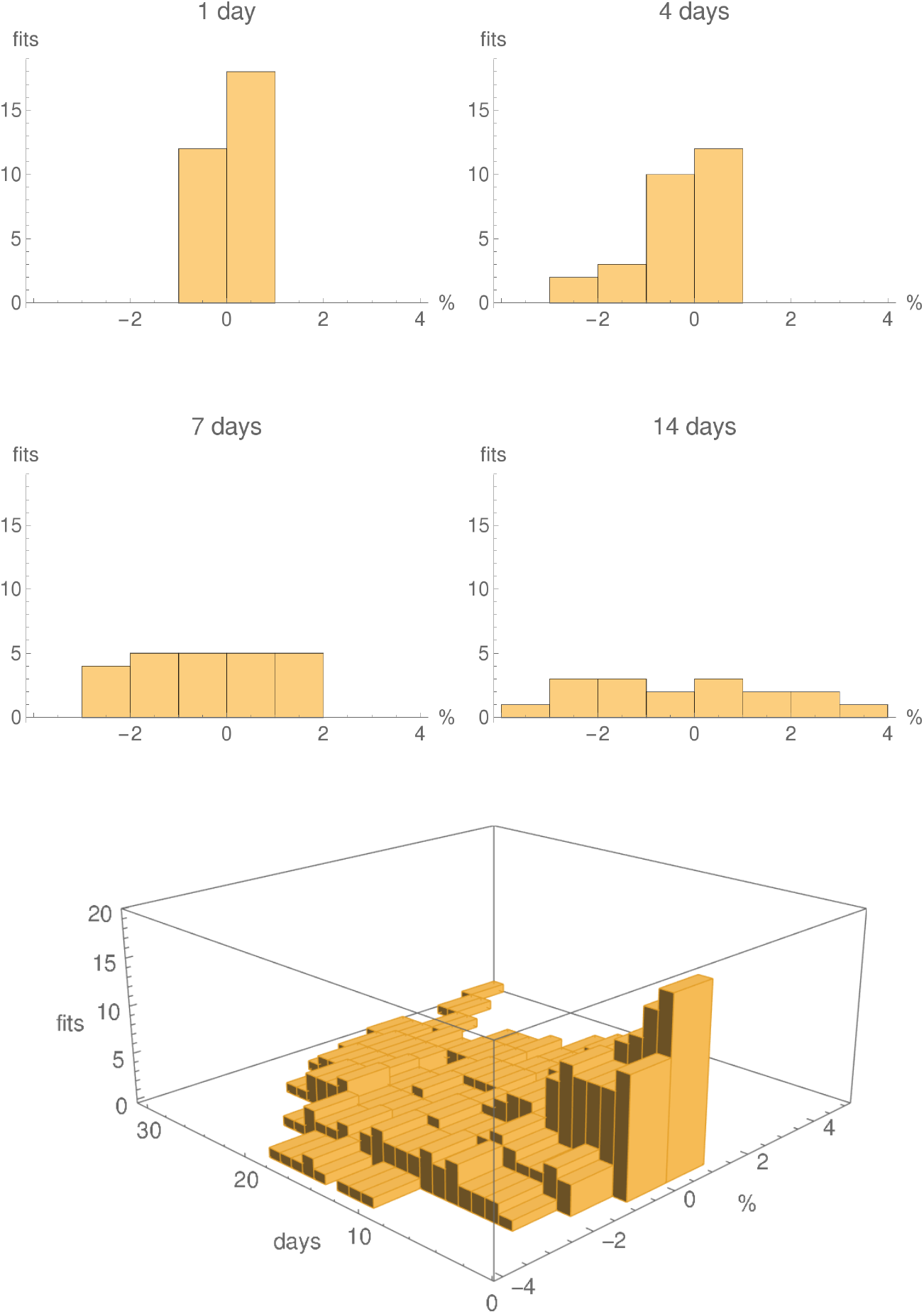
Histograms showing the numbers predictions with 1% error bins for the logistic fits with running parameters, for Italy, generated in the period April 5 - May 4, for 1 days, 4 days, one week and two weeks. The last row shows a 3D histogram for all 30 fits. The individual histograms in the first two rows are slices, at the given number of days, of the 3D histogram.

**Figure A.7:**
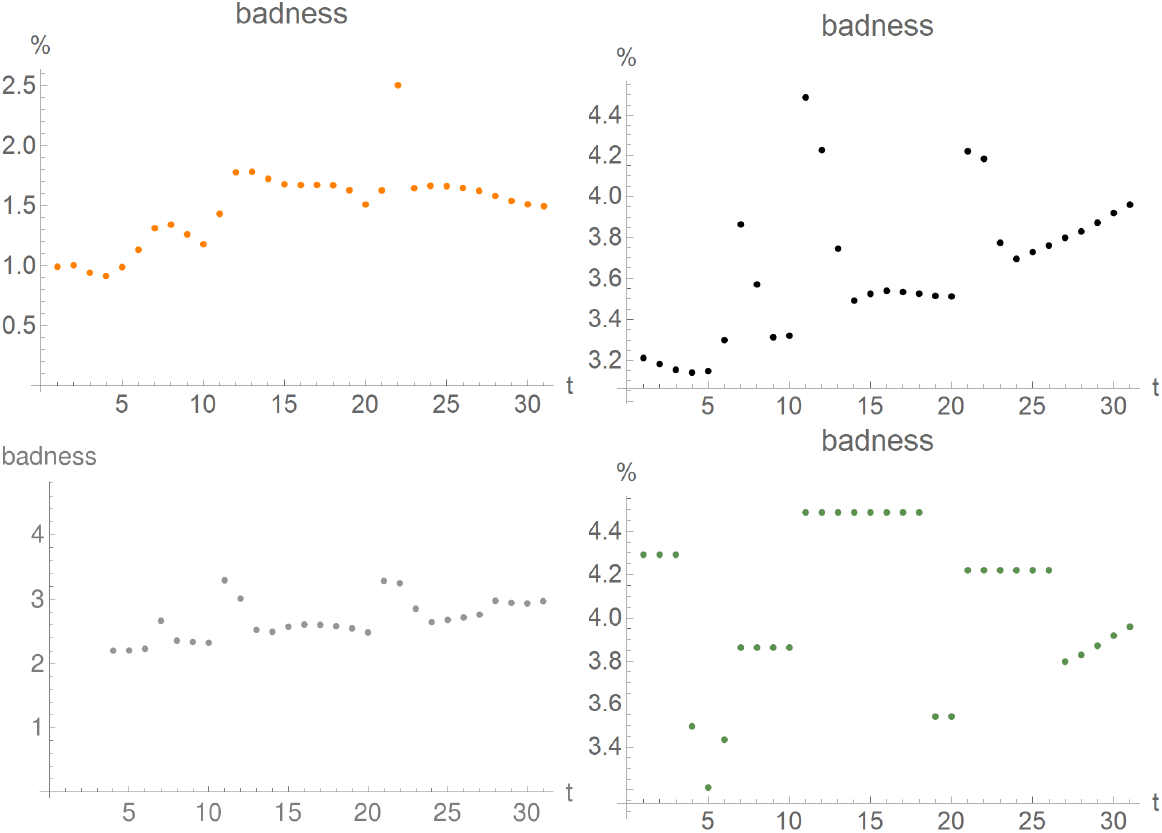
First “badness parameter” for fits for New York generated in the period April 5 (day 1 on the plot) - May 5 (day 31 on the plot). Upper row: double logistic (left), logistic (right); lower row: Gauss-error (left), logistic with running constants (right).

**Figure A.8:**
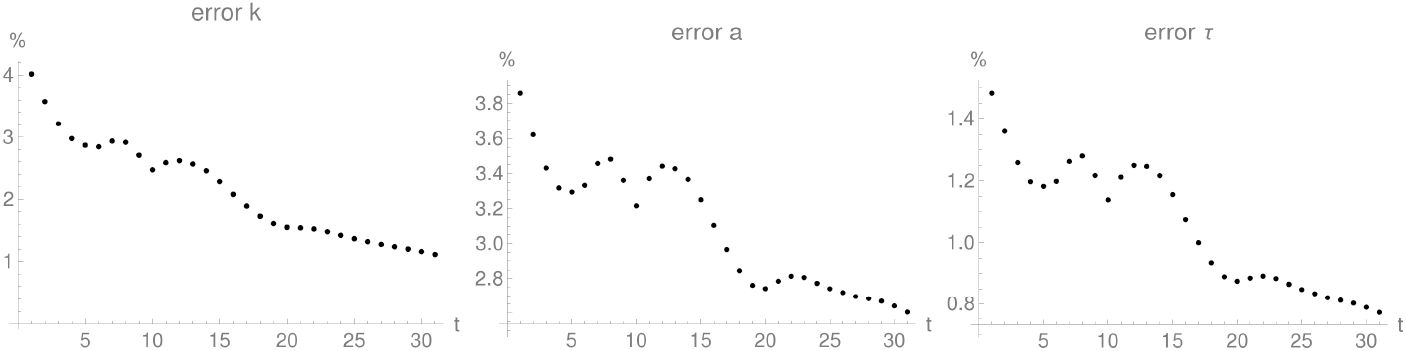
Relative error of logistic-fit parameters for New York generated in the period April 5 (day 1 on the plot) - May 5 (day 31 on the plot).

### A.2 New York

In this section we present the results of our analysis for New York, in parallel to the presentation of Austria in Section 5.

One would have avoided the worst predictions by deciding, after the first week of fitting, to reject those predictions where the second badness parameter is larger than 7%.

The plots of the first badness parameter are found in Figure A.7, while the relative errors of the fits can be found in Figure A.8.

The fits with their standard errors and residuals are found in Figure A.9.

In Figure A.10 we show how the parameters of the fits, and their confidence intervals, change in time, for the double logistic, Gauss-error, and logistic curves.

The predictability strings for New York are plotted in Figure A.11.

In Figure A.12 we show the histograms of the errors of the predictions.

**Figure A.9:**
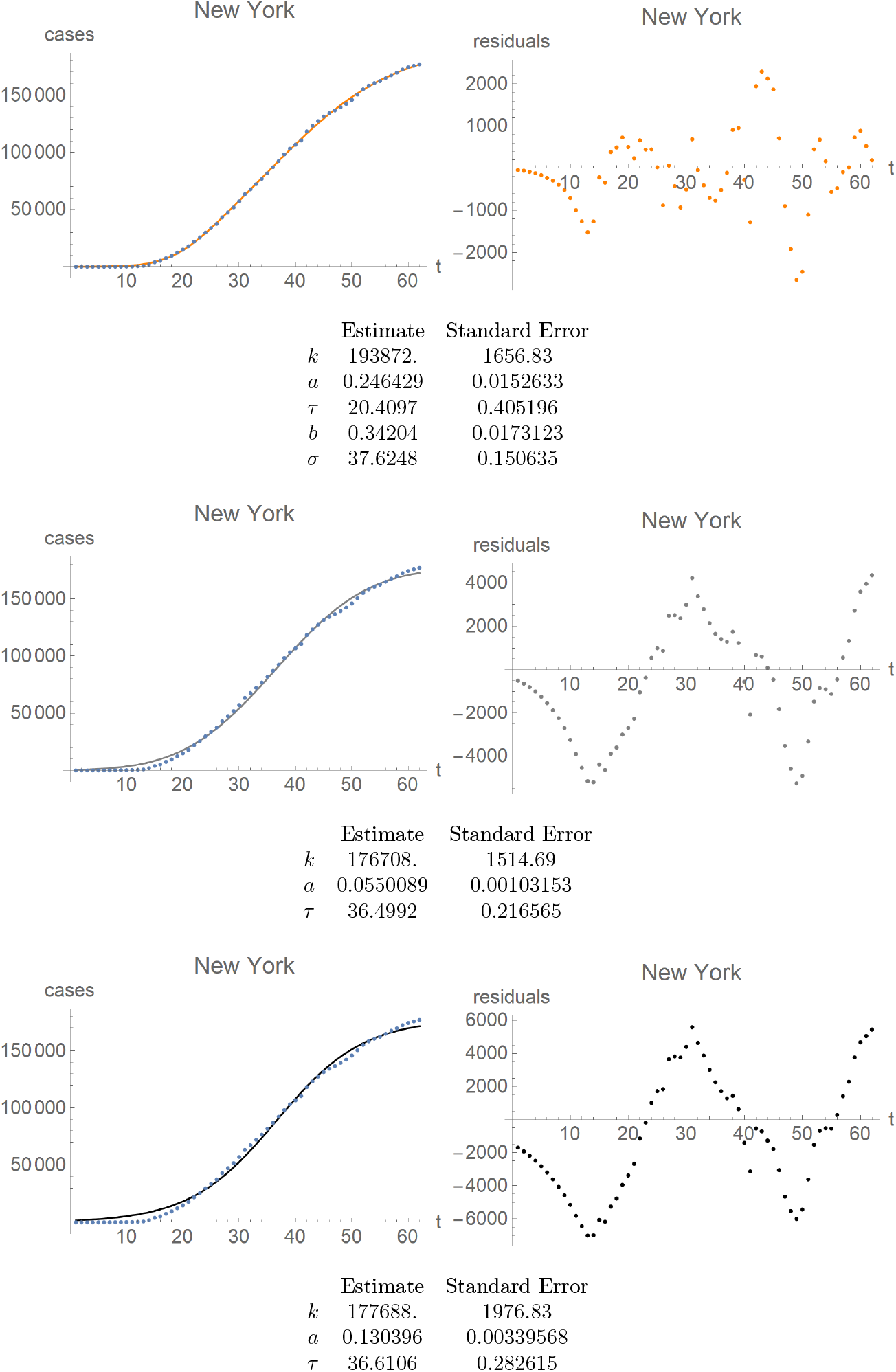
New York. First row: fit of the double-logistic curve (left) and its residuals (right). Same for the Gauss-error curve in the second row, and the logistic curves in the third row. Note distinct scales for the residual plots as compared to the fitted curve, as well as a distinct scale for the residuals of the double-logistic curve compared to the other residuals.

**Figure A.10:**
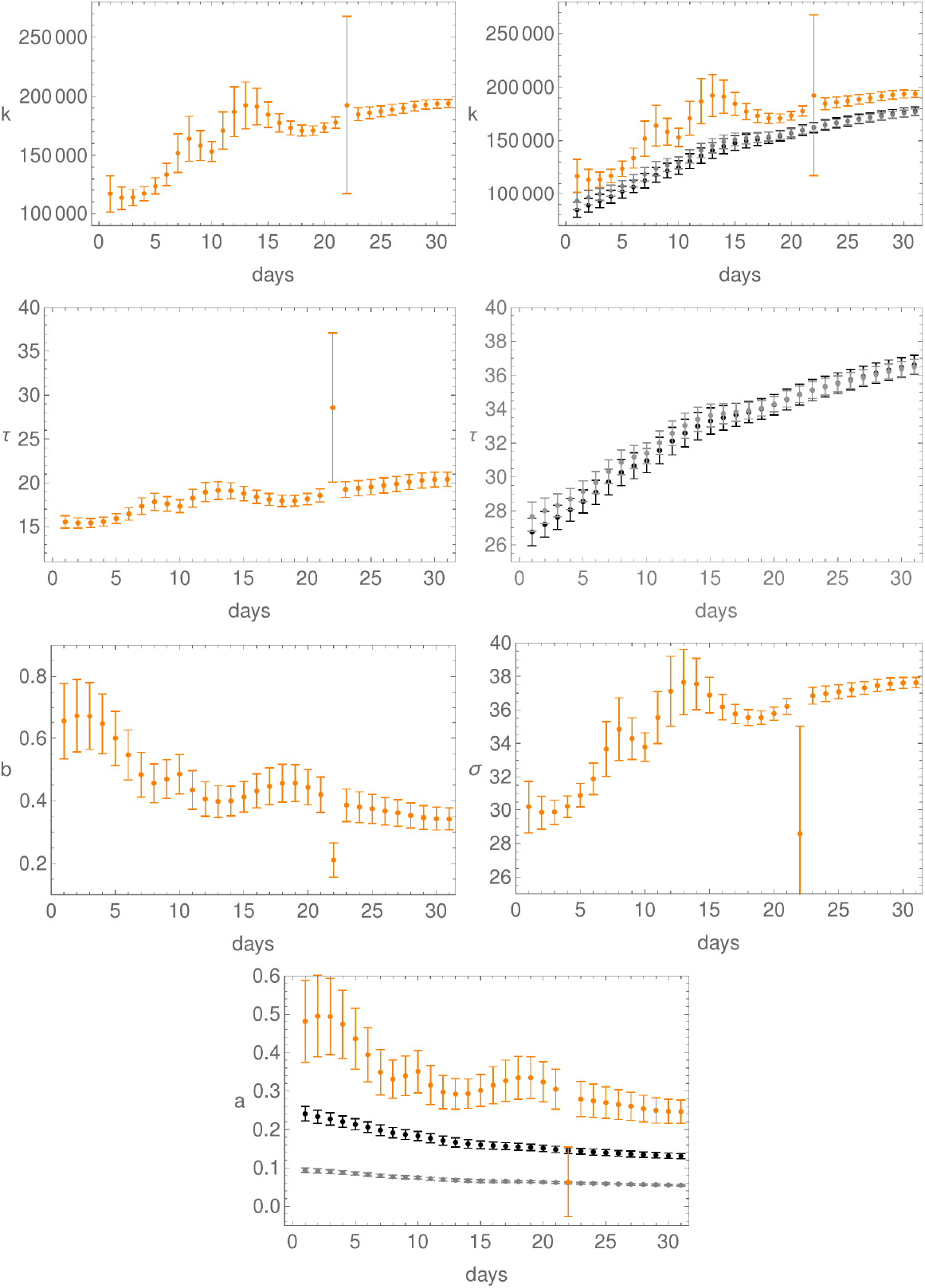
Evolution of the fit parameters in time, together with their confidence intervals, for New York; day 1 is April 5. First row: parameter *k* for the double-logistic curve (left), and for all three curves (right). Here, as elsewhere, orange is for the double-logistic, black for the logistic, and grey for the Gauss-error function. Second row: the *τ*-parameter for the double-logistic curve (left) and for both the logistic and Gauss-error curves (right). Third row: plots of the parameters *b* (left) and *σ* (right) for the double-logistic curve. Last row: plot of the parameter *a* for all three curves.

**Figure A.11:**
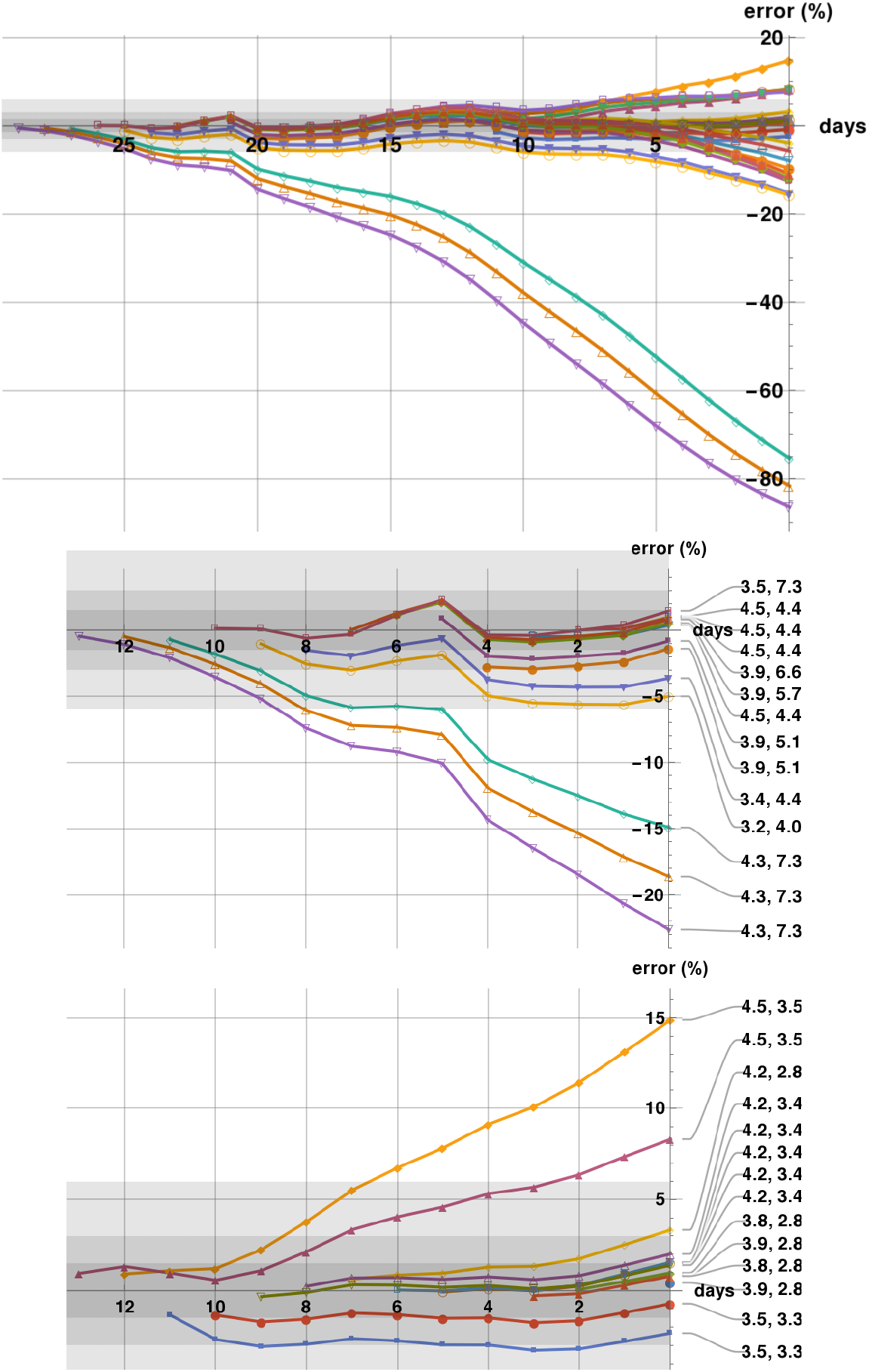
Predictability plots for New York generated in the period April 5 - May 4. The first row shows all predictability strings with last prediction on May 5, which is the last day on the first and third rows. The shadings correspond to a 1.5%, 3%, and 6% error zones. The second row shows the predictability strings cut-off at April 20, with badness parameters indicated. The third row shows only these predictability strings which start within a two-weeks period ending on May 5.

**Figure A.12:**
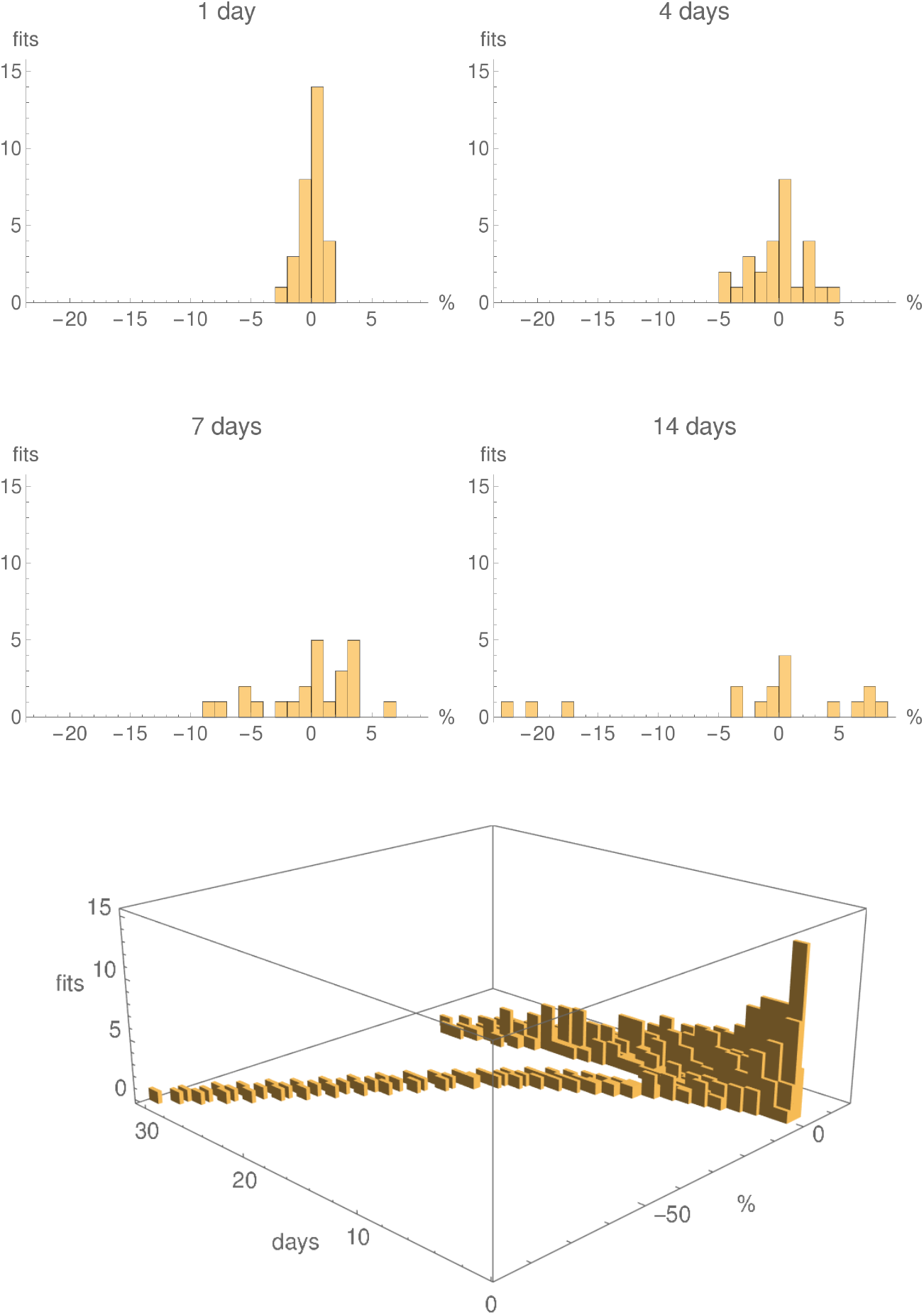
Histograms showing the numbers predictions with 1% error bins for the logistic fits with running parameters, for New York, generated in the period April 5 - May 4, for 1 days, 4 days, one week and two weeks. The last row shows a 3D histogram for all 30 fits. The individual histograms in the first two rows are slices, at the given number of days, of the 3D histogram.

**Figure A.13:**
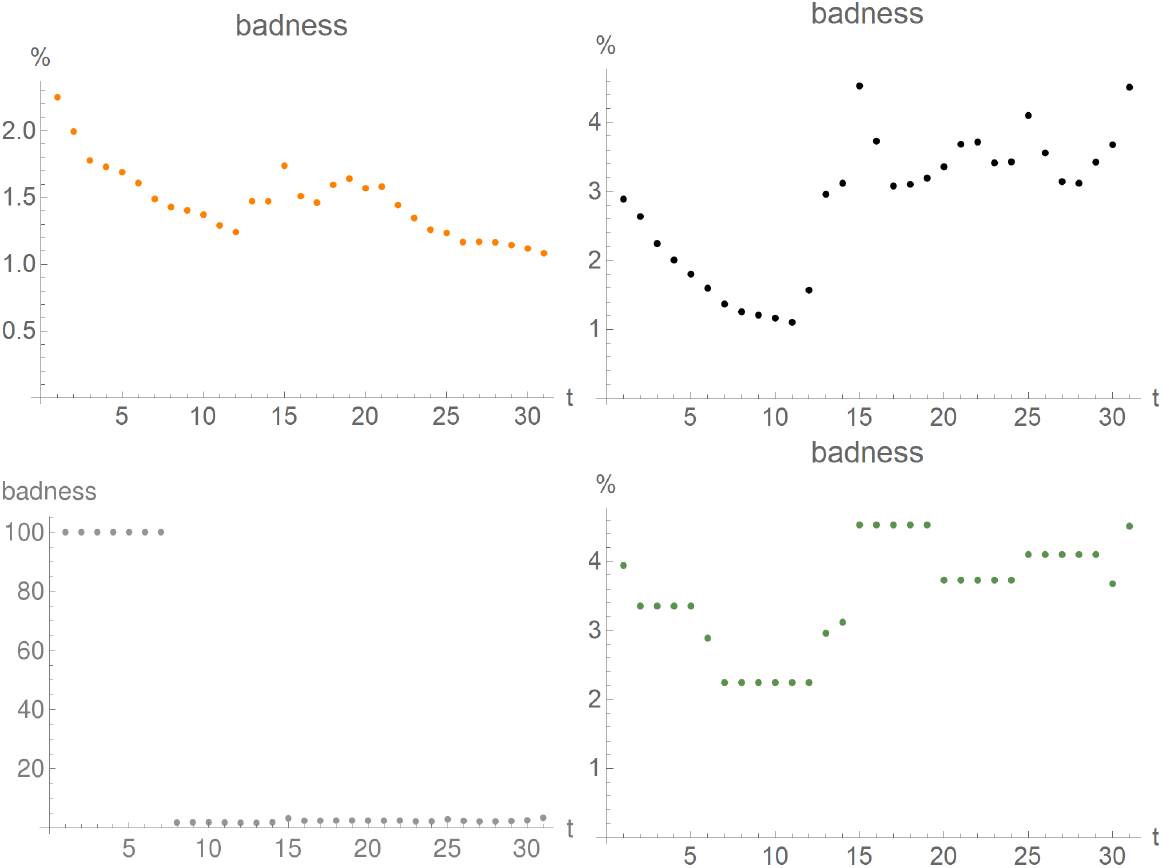
First “badness parameter” for fits for Poland generated in the period April 5 (day 1 on the plot) - May 5 (day 31 on the plot). Upper row: double logistic (left), logistic (right); lower row: Gauss-error (left), logistic with running constants (right).

**Figure A.14:**
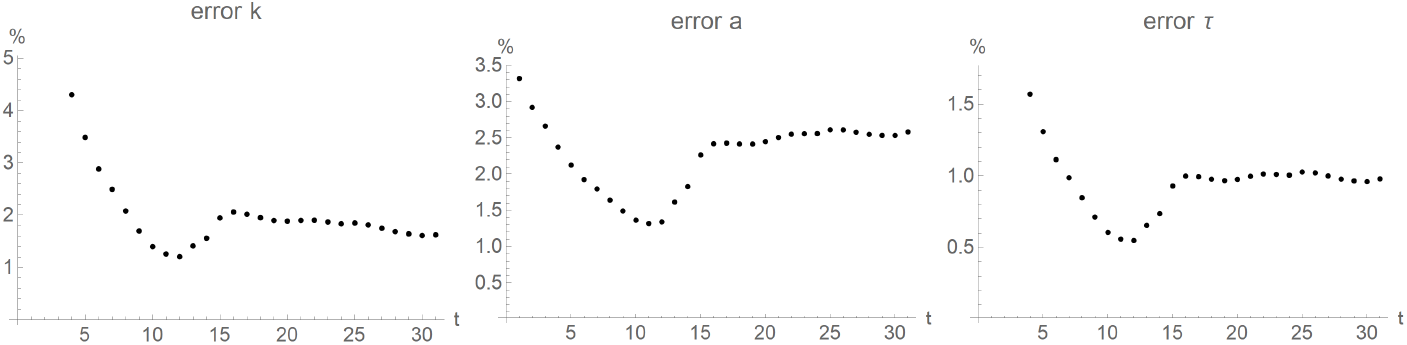
Relative error of logistic-fit parameters for Poland generated in the period April 5 (day 1 on the plot) - May 5 (day 31 on the plot).

### A.3 Poland

In this section we present the results of our analysis for Poland, in parallel to the presentation of Austria in Section 5.

Similarly to New York, one would have avoided the worst predictions by deciding, after the first week of fitting, to reject those predictions where the second badness parameter is larger than 7%.

The plots of the first badness parameter are found in Figure A.13, while the relative errors of the fits can be found in Figure A.14.

The fits with their standard errors and residuals are found in Figure A.15.

In Figure A.16 we show how the parameters of the fits, and their confidence intervals, change in time, for the double logistic, Gauss-error, and logistic curves.

The predictability strings for Poland are plotted in Figure A.17.

In Figure A.18 we show the histograms of the errors of the predictions.Poland residuals

**Figure A.15:**
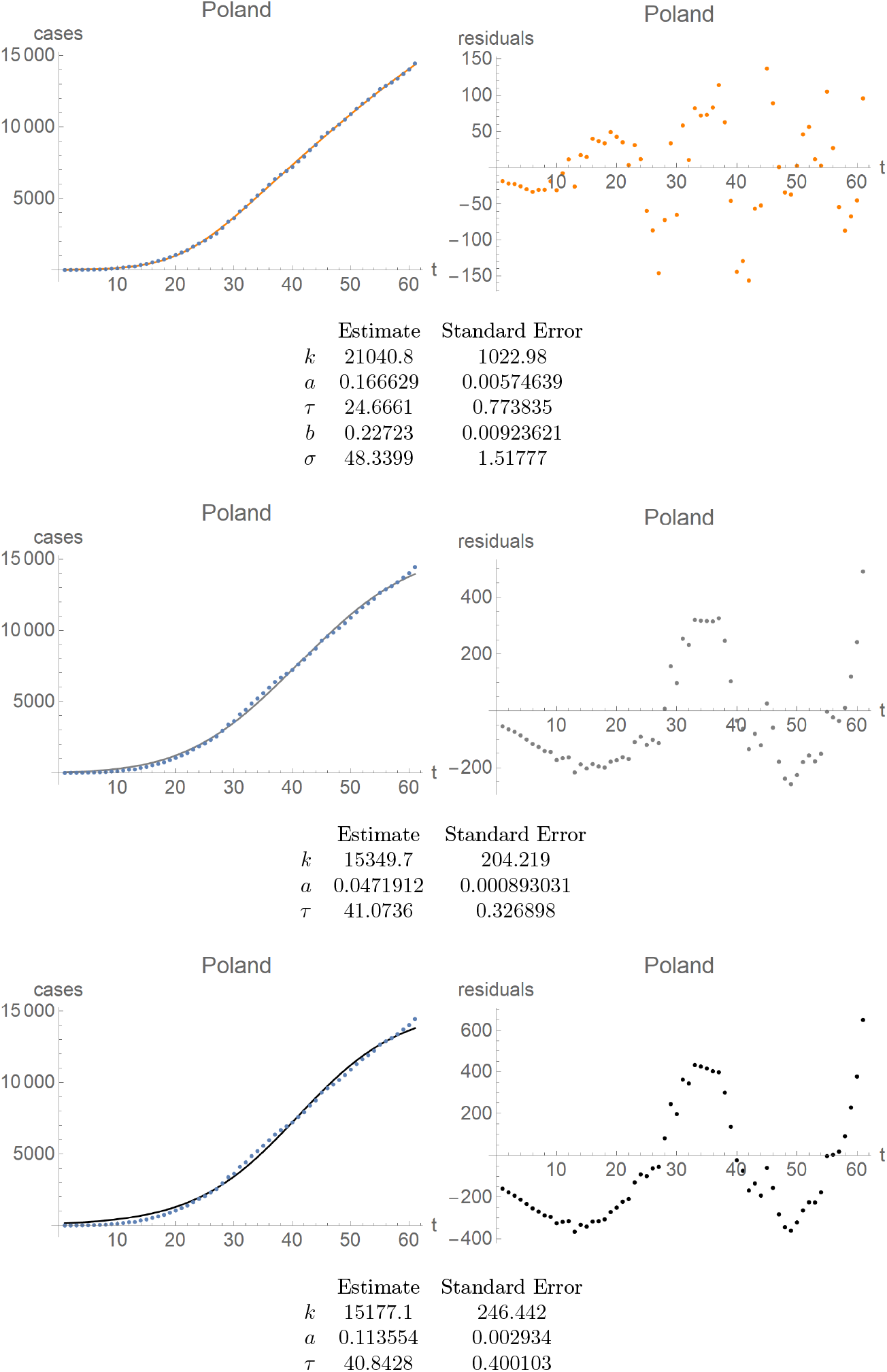
Poland. First row: fit of the double-logistic curve (left) and its residuals (right). Same for the Gauss-error curve in the second row, and the logistic curves in the third row. Note distinct scales for the residual plots as compared to the fitted curve, as well as a distinct scale for the residuals of the double-logistic curve compared to the other residuals.

**Figure A.16:**
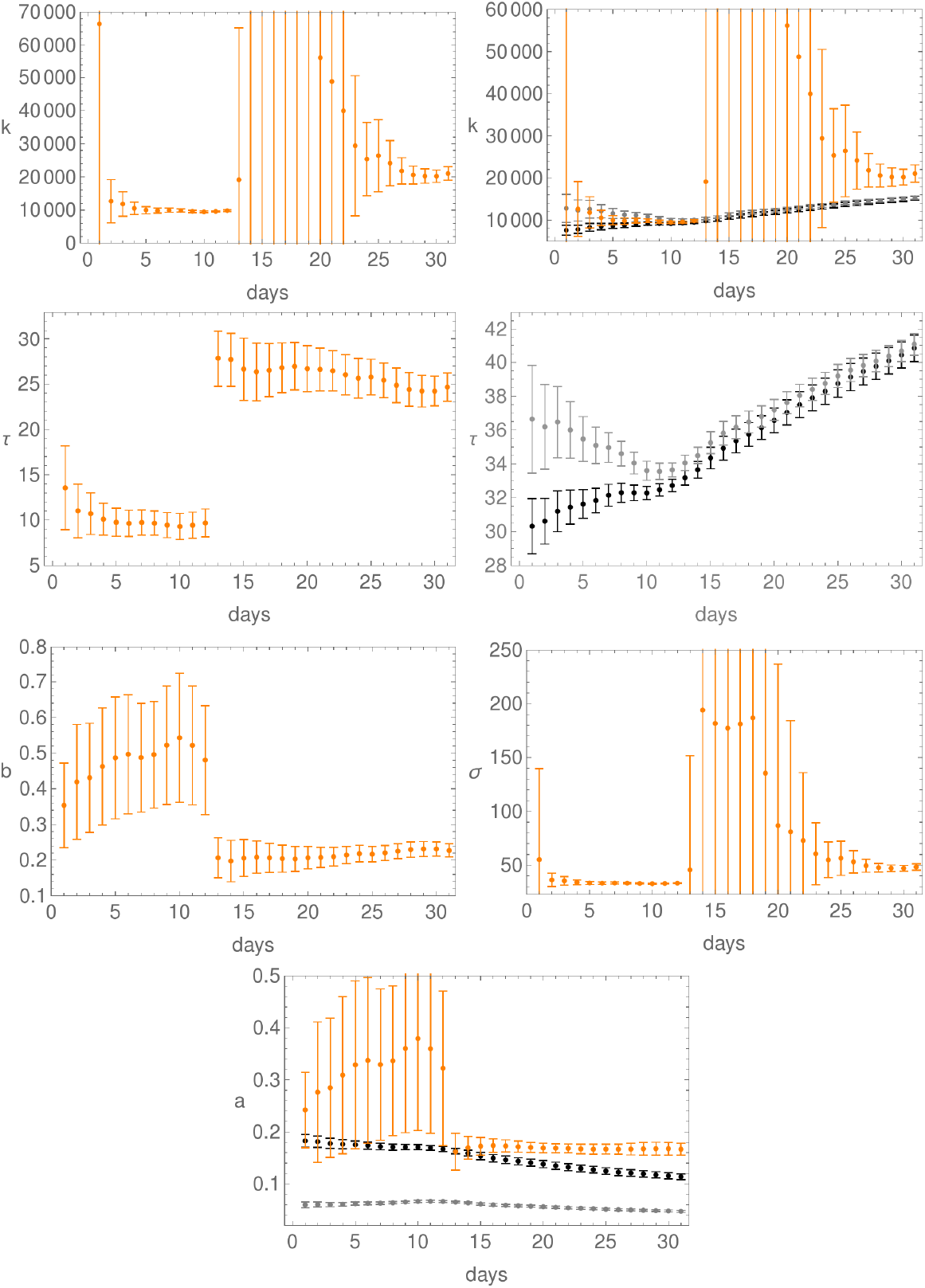
Evolution of the fit parameters in time, together with their confidence intervals, for Poland; day 1 is April 5. First row: parameter *k* for the double-logistic curve (left), and for all three curves (right). Here, as elsewhere, orange is for the double-logistic, black for the logistic, and grey for the Gauss-error function. Second row: the *τ*-parameter for the double-logistic curve (left) and for both the logistic and Gauss-error curves (right). Third row: plots of the parameters *b* (left) and *σ* (right) for the double-logistic curve. Last row: plot of the parameter *a* for all three curves.

**Figure A.17:**
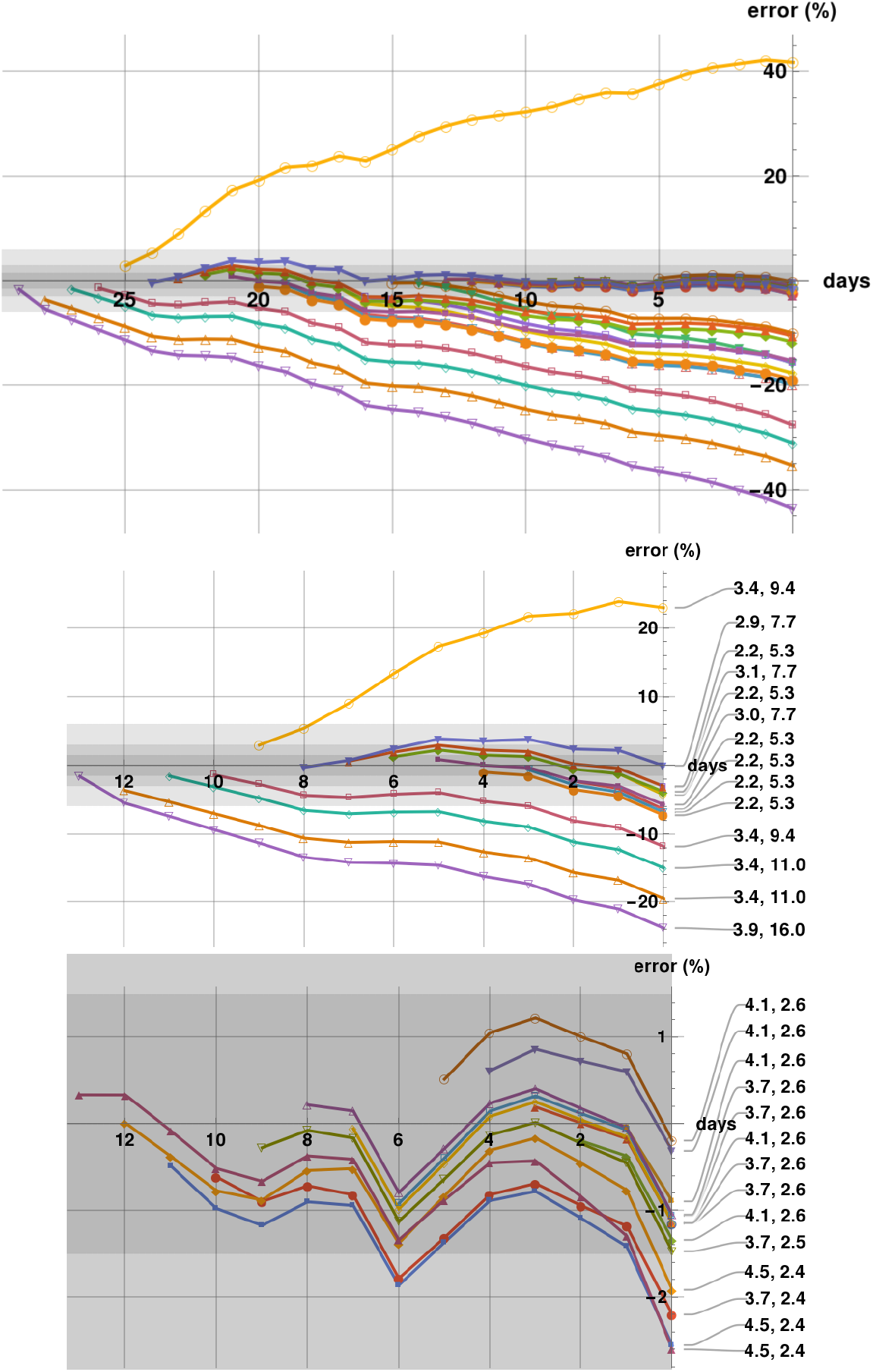
Predictability plots for Poland generated in the period April 5 - May 4. The first row shows all predictability strings with last prediction on May 5, which is the last day on the first and third rows. The shadings correspond to a 1.5%, 3%, and 6% error zones. The second row shows the predictability strings cut-off at April 20, with badness parameters indicated. The third row shows only these predictability strings which start within a two-weeks period ending on May 5.

**Figure A.18:**
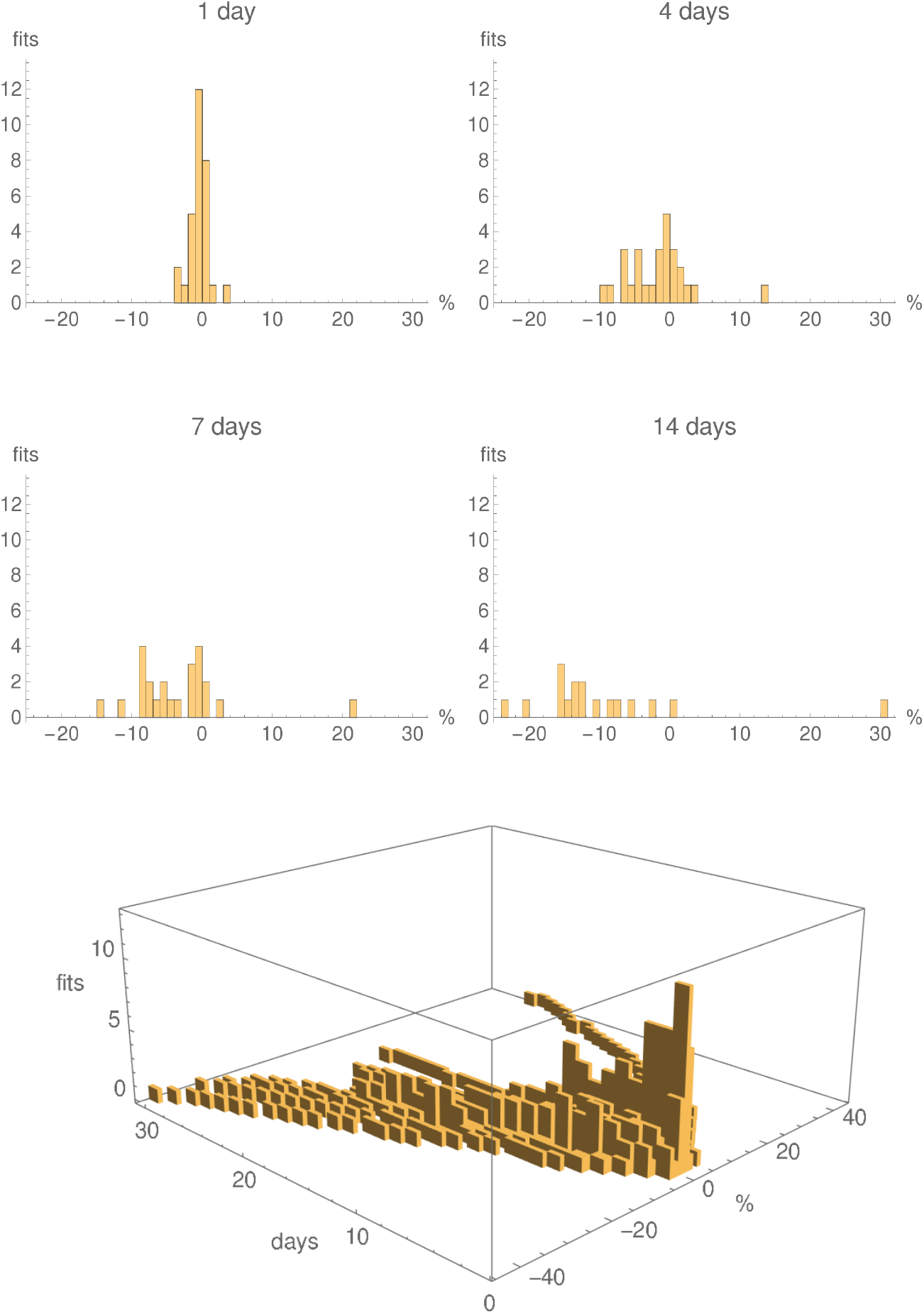
Histograms showing the numbers predictions with 1% error bins for the logistic fits with running parameters, for Poland, generated in the period April 5 - May 4, for 1 days, 4 days, one week and two weeks. The last row shows a 3D histogram for all 30 fits. The individual histograms in the first two rows are slices, at the given number of days, of the 3D histogram.

**Figure A.19:**
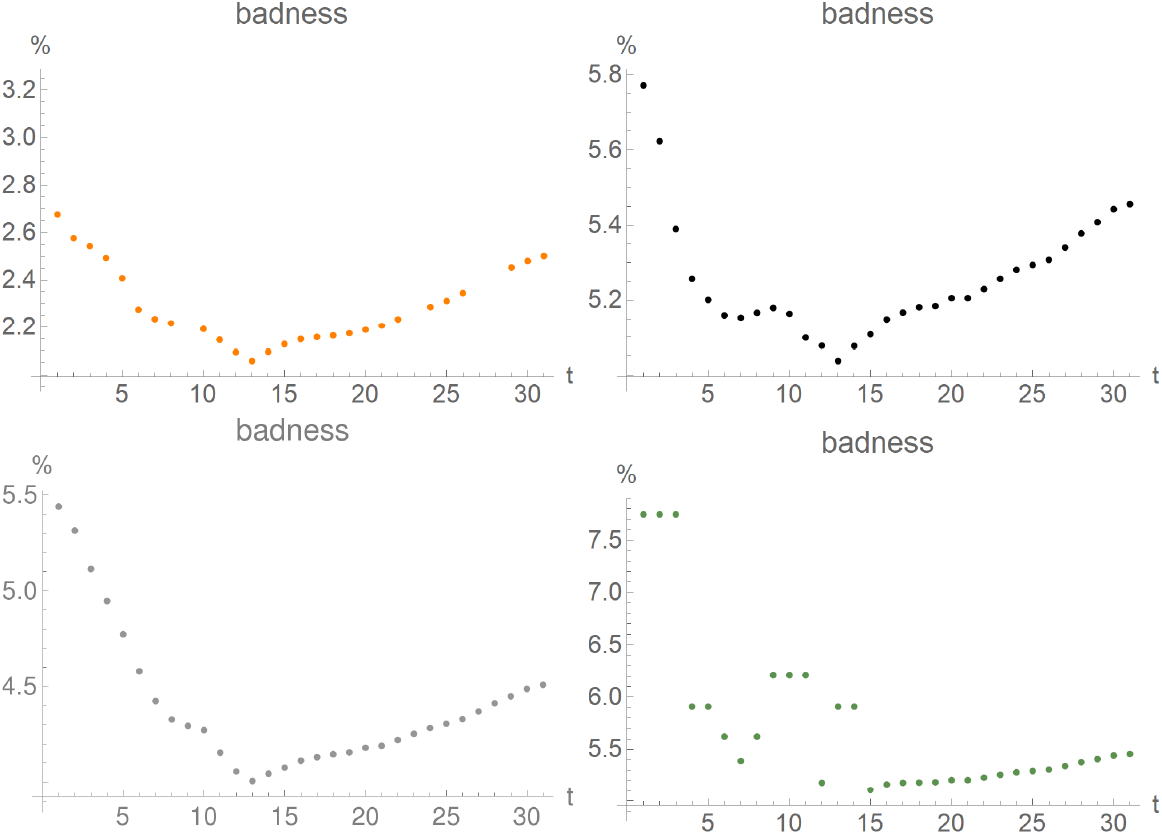
First “badness parameter” for fits for Slovenia generated in the period April 5 (day 1 on the plot) - May 5 (day 31 on the plot). Upper row: double logistic (left), logistic (right); lower row: Gauss-error (left), logistic with running constants (right).

**Figure A.20:**
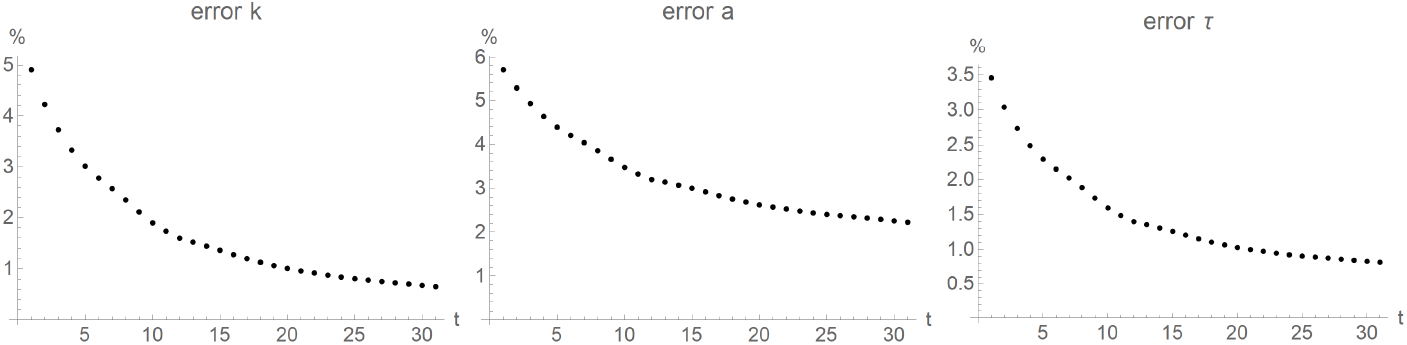
Relative error of logistic-fit parameters for Slovenia generated in the period April 5 (day 1 on the plot) - May 5 (day 31 on the plot).

### A.4 Slovenia

We included Slovenia in our analysis as it is one of the first countries to suspend all confinement measures, on May 15, 2020. All residuals are smaller than about 2% of the total number of cases; and smaller than about 1.6% when two worse residuals are excluded. The number of cases is about 1/10th of that of Austria, but the double-logistic curve has distinctively different parameters. On the other had, its residuals are again of the order of 1/10th of those of Austria. The predictability algorithm which excludes predictions with badness parameters larger than 7% would have produced rather good predictions for the whole month of the epidemics under consideration.

The plots of the first badness parameter are found in Figure A.19, while the relative errors of the fits can be found in Figure A.20.

The fits with their standard errors and residuals are found in Figure A.21.

**Figure A.21:**
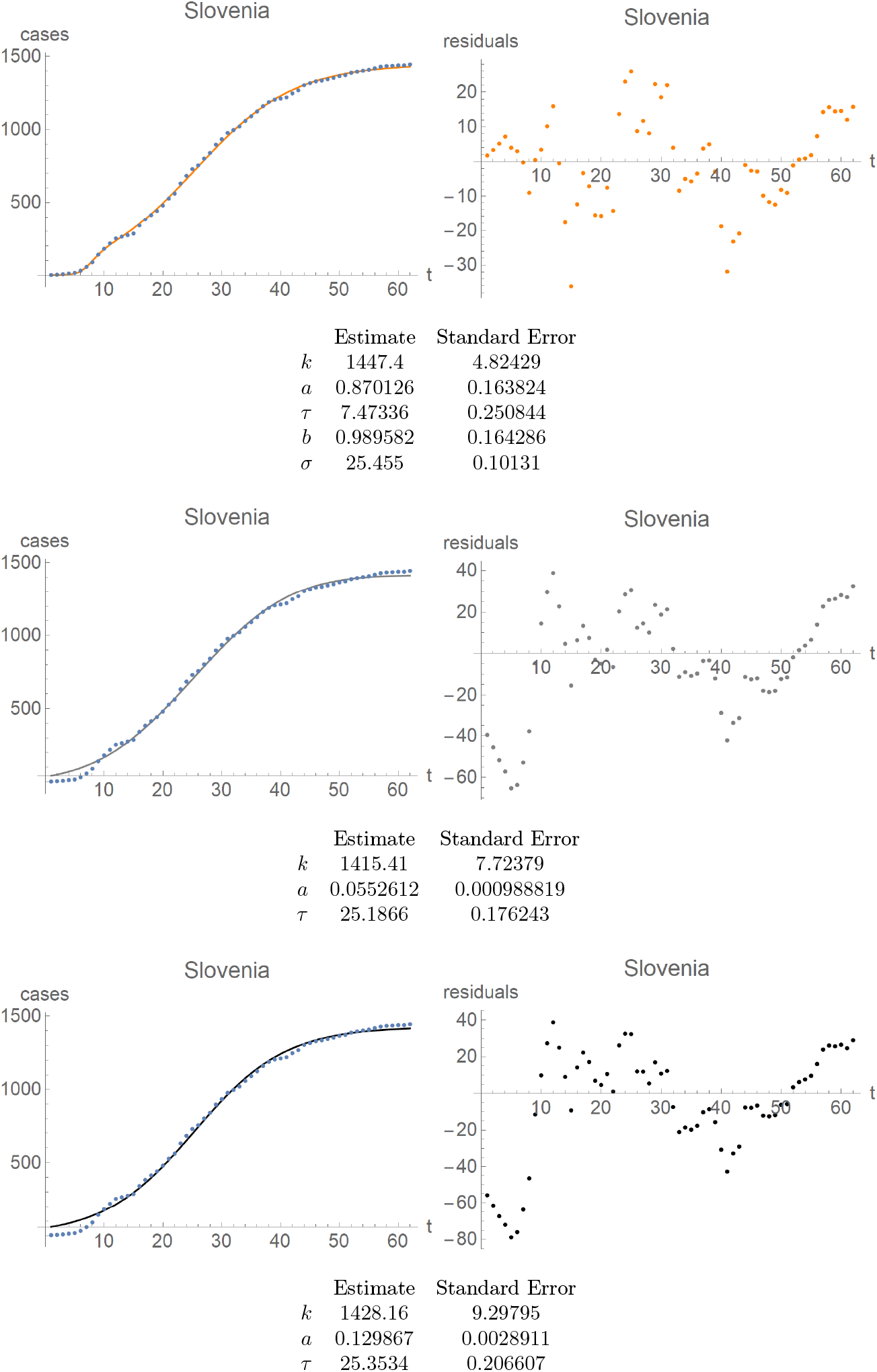
Slovenia. First row: fit of the double-logistic curve (left) and its residuals (right). Same for the Gauss-error curve in the second row, and the logistic curves in the third row. Note distinct scales for the residual plots as compared to the fitted curve, as well as a distinct scale for the residuals of the double-logistic curve compared to the other residuals.

In Figure A.22 we show how the parameters of the fits, and their confidence intervals, change in time, for the double logistic, Gauss-error, and logistic curves.

The predictability strings for Slovenia are plotted in Figure A.23.

In Figure A.24 we show the histograms of the errors of the predictions.

**Figure A.22:**
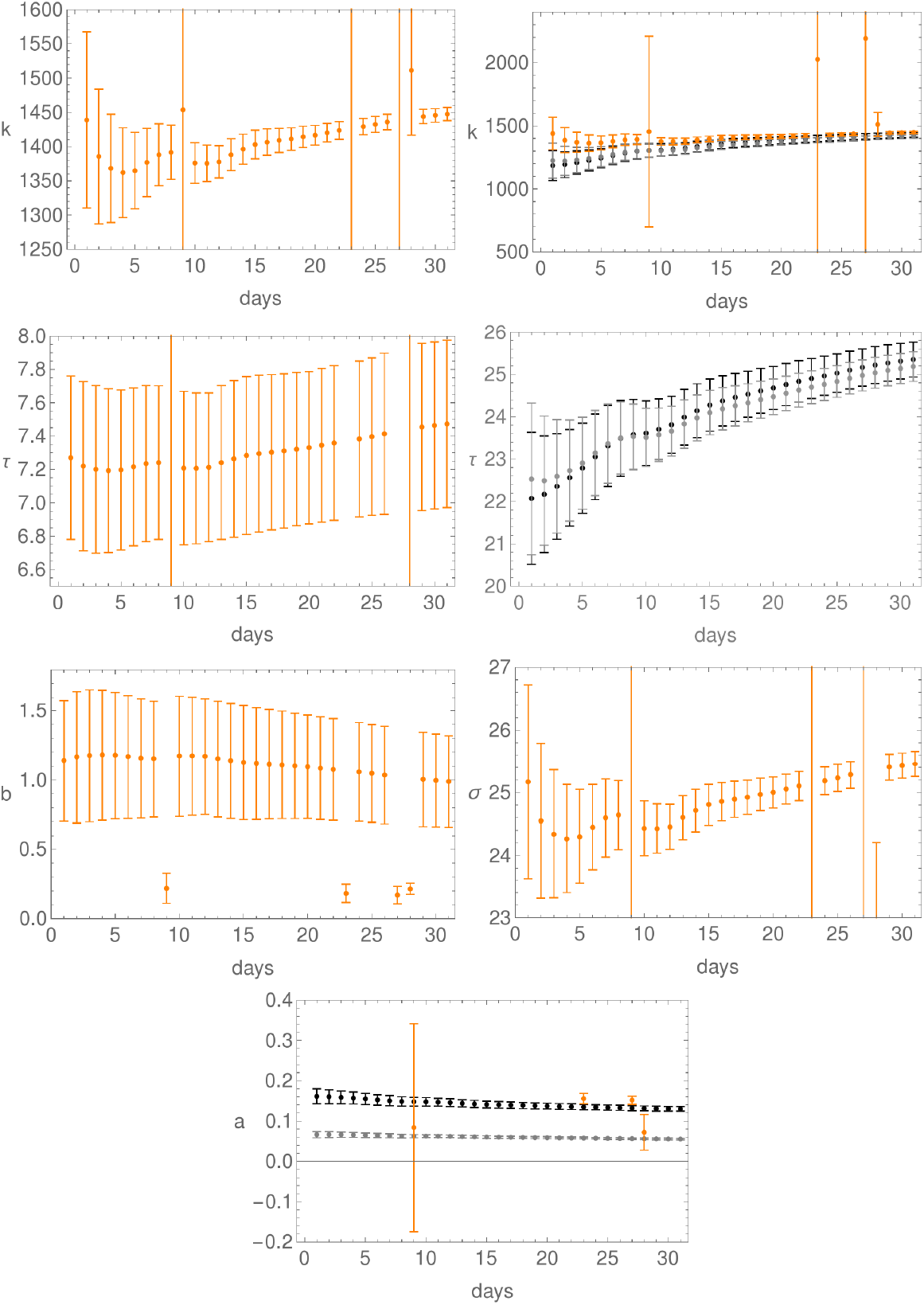
Evolution of the fit parameters in time, together with their confidence intervals, for Slovenia; day 1 is April 5. First row: parameter *k* for the double-logistic curve (left), and for all three curves (right). Here, as elsewhere, orange is for the double-logistic, black for the logistic, and grey for the Gauss-error function. Second row: the *τ*-parameter for the double-logistic curve (left) and for both the logistic and Gauss-error curves (right). Third row: plots of the parameters *b* (left) and *σ* (right) for the double-logistic curve. Last row: plot of the parameter *a* for all three curves.

**Figure A.23:**
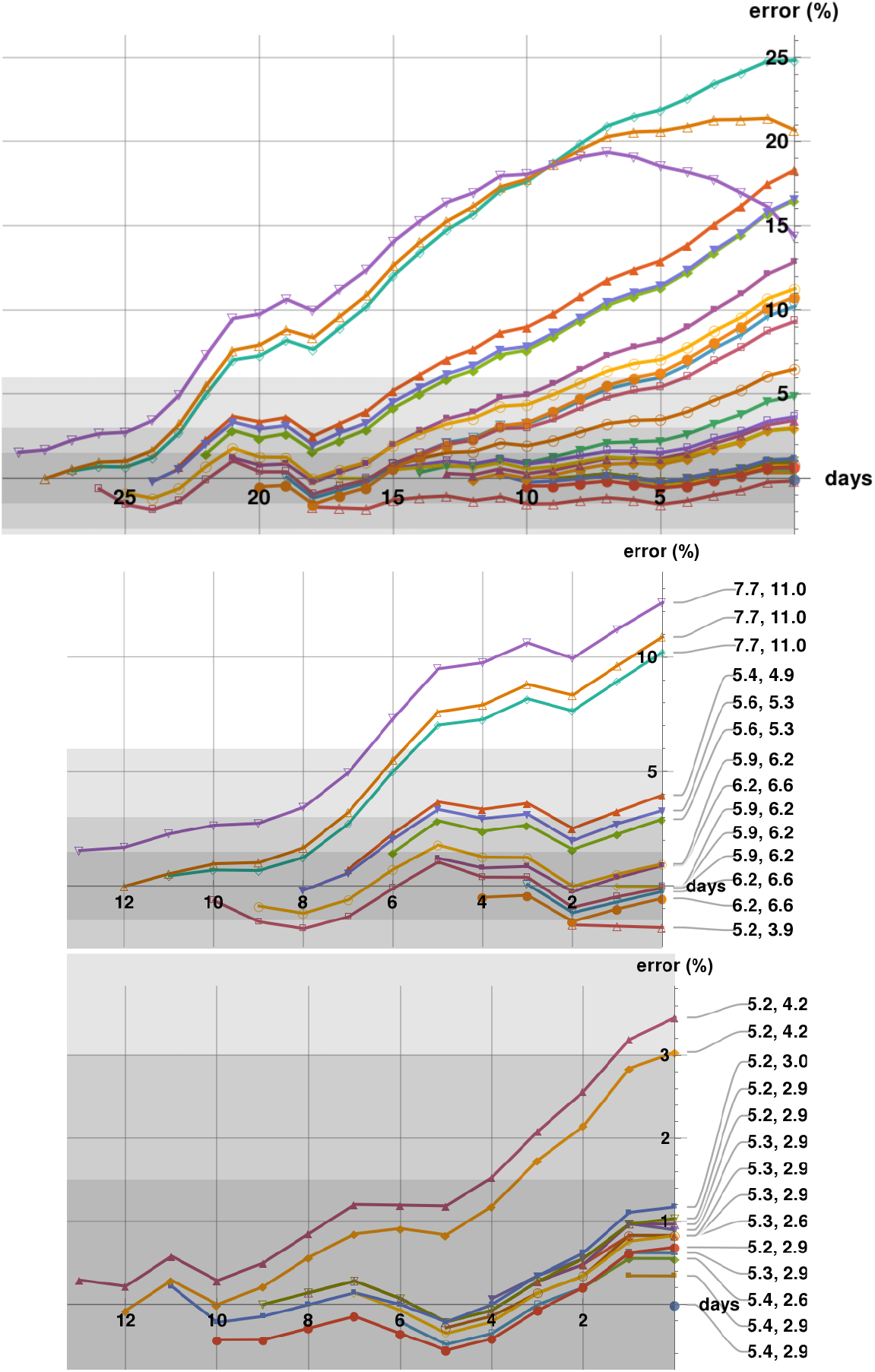
Predictability plots for Slovenia generated in the period April 5 - May 4. The first row shows all predictability strings with last prediction on May 5, which is the last day on the first and third rows. The shadings correspond to a 1.5%, 3%, and 6% error zones. The second row shows the predictability strings cut-off at April 20, with badness parameters indicated. The third row shows only these predictability strings which start within a two-weeks period ending on May 5.

**Figure A.24:**
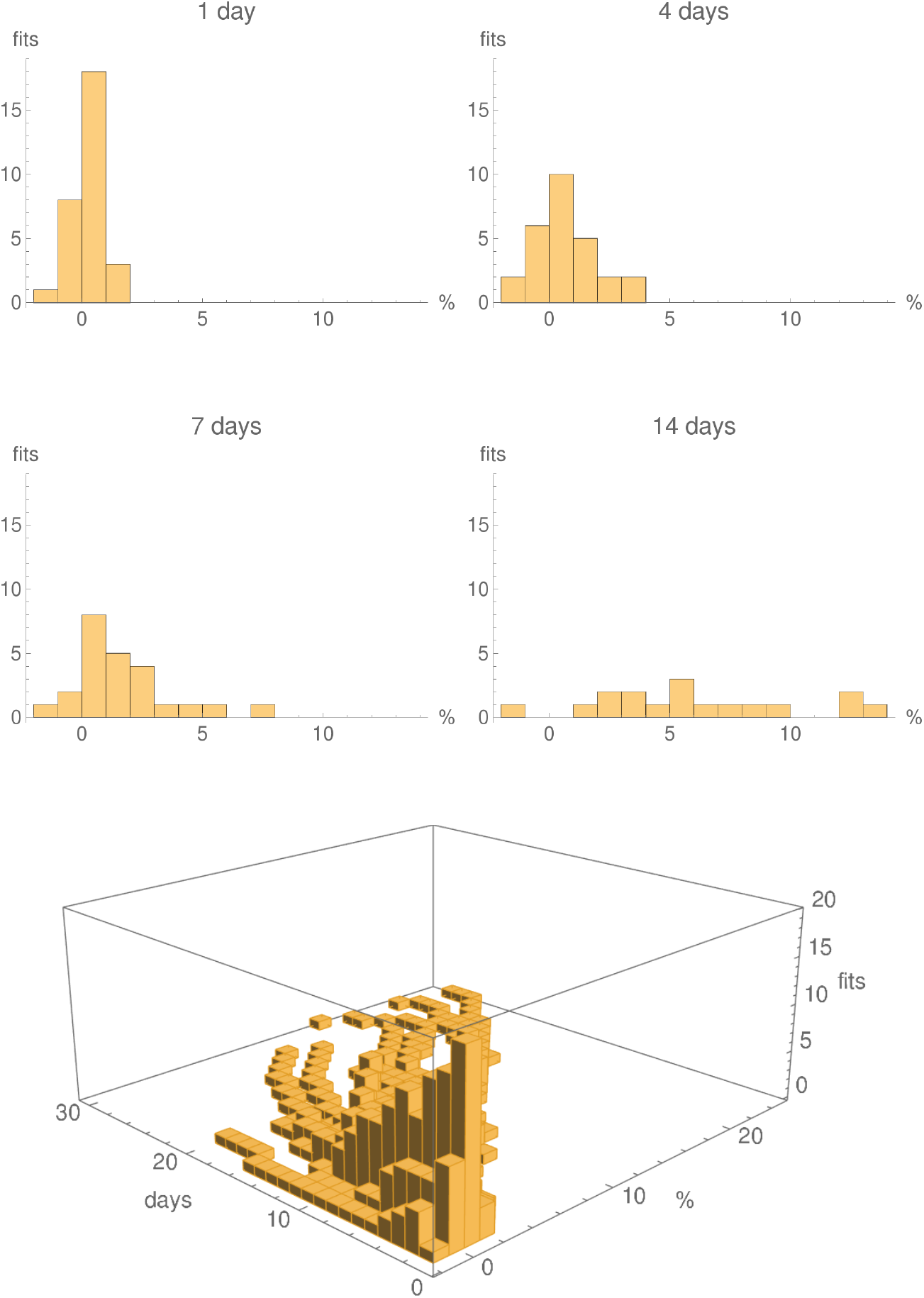
Histograms showing the numbers predictions with 1% error bins for the logistic fits with running parameters, for Slovenia, generated in the period April 5 - May 4, for 1 days, 4 days, one week and two weeks. The last row shows a 3D histogram for all 30 fits. The individual histograms in the first two rows are slices, at the given number of days, of the 3D histogram.

**Figure A.25:**
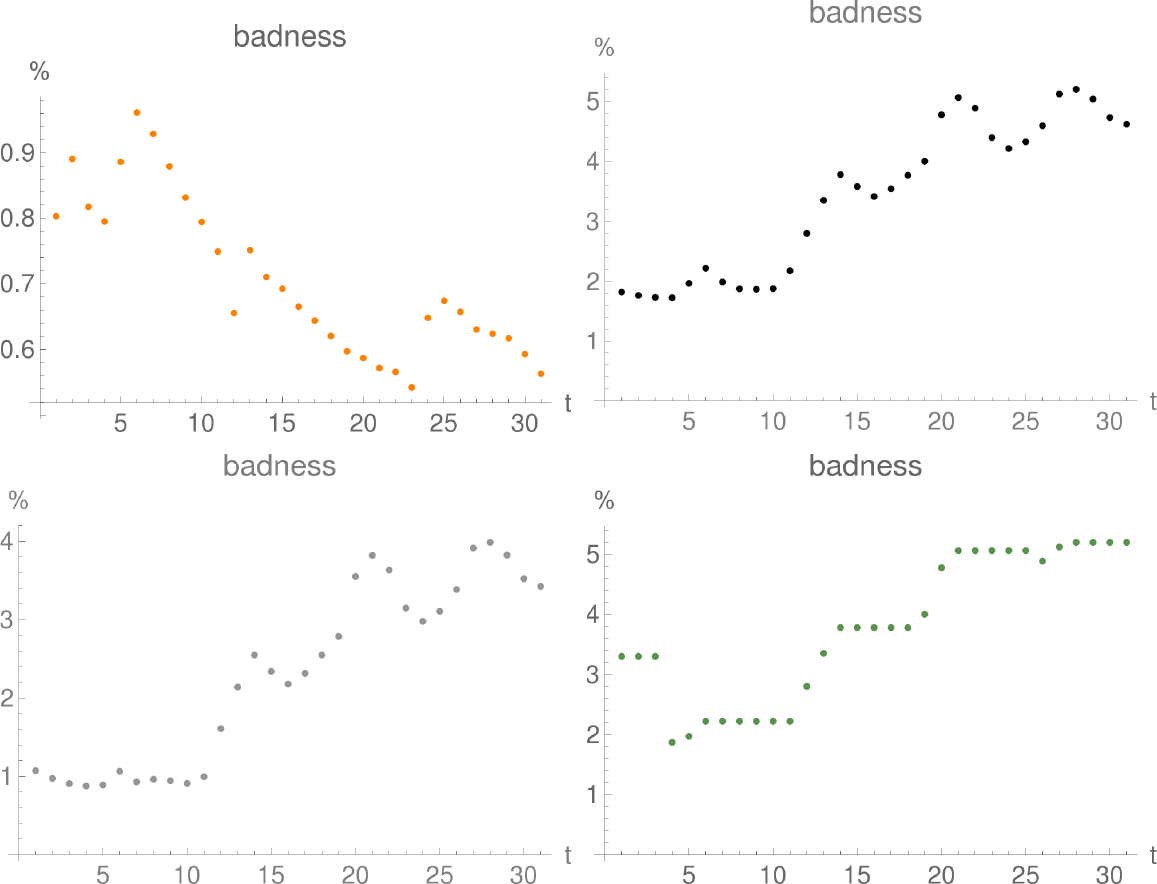
First “badness parameter” for fits for US generated in the period April 5 (day 1 on the plot) - May 5 (day 31 on the plot). Upper row: double logistic (left), logistic (right); lower row: Gauss-error (left), logistic with running constants (right).

### A.5 US

In this section we present the results of our analysis for the US, in parallel to the presentation of Austria in Section 5.

To avoid the worst predictions one should have decided, after the first week of fitting, to reject those predictions where the second badness parameter is larger than 5%. Note that the 7% threshold, seen in other countries, would not have worked here.

The plots of the first badness parameter are found in Figure A.25, while the relative errors of the fits can be found in Figure A.26.

The fits with their standard errors and residuals are found in Figure A.27.

**Figure A.26:**
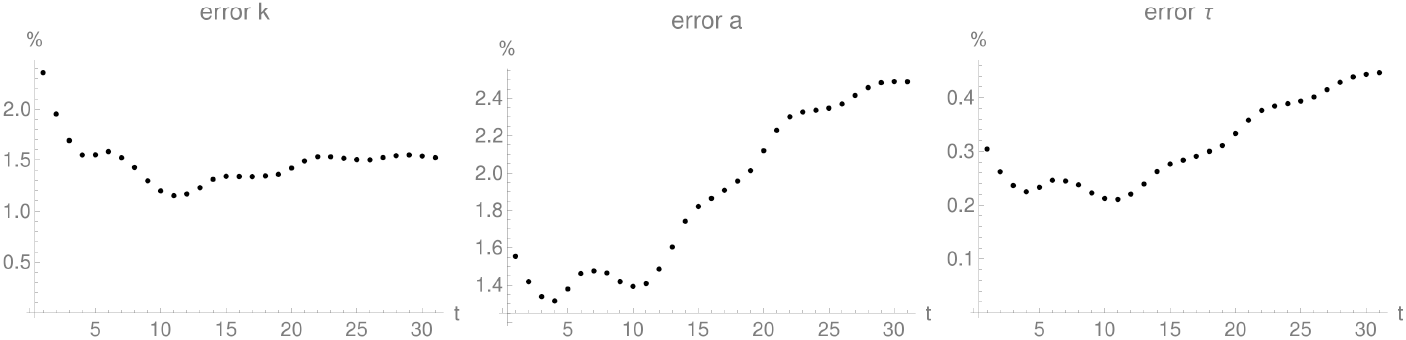
Relative error of logistic-fit parameters for US generated in the period April 5 (day 1 on the plot) - May 5 (day 31 on the plot).

**Figure A.27:**
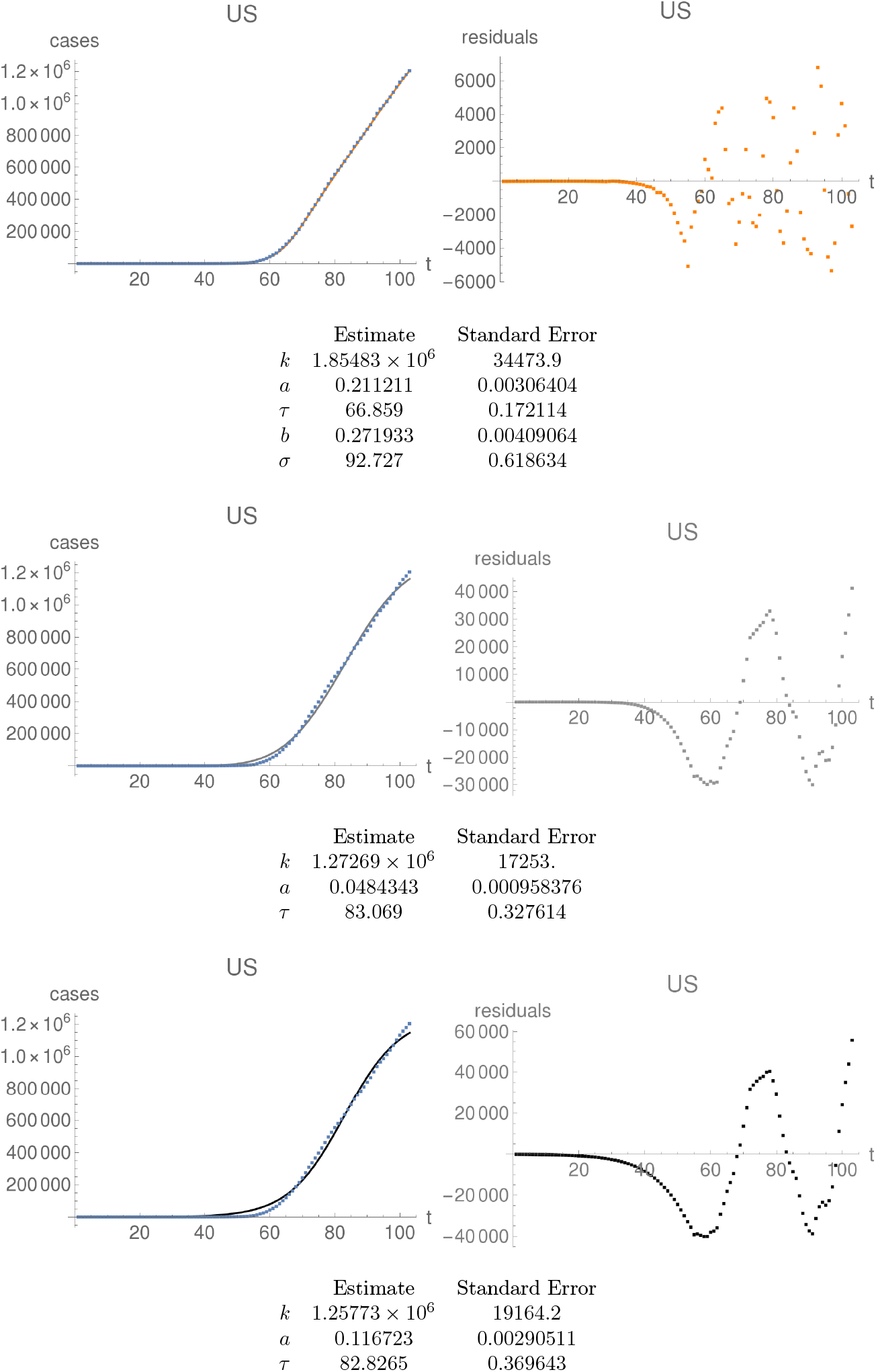
US. First row: fit of the double-logistic curve (left) and its residuals (right). Same for the Gauss-error curve in the second row, and the logistic curves in the third row. Note distinct scales for the residual plots as compared to the fitted curve, as well as a distinct scale for the residuals of the double-logistic curve compared to the other residuals.

In Figure A.28 we show how the parameters of the fits, and their confidence intervals, change in time, for the double logistic, Gauss-error, and logistic curves.

The predictability strings for US are plotted in Figure A.29.

In Figure A.30 we show the histograms of the errors of the predictions.

**Figure A.28:**
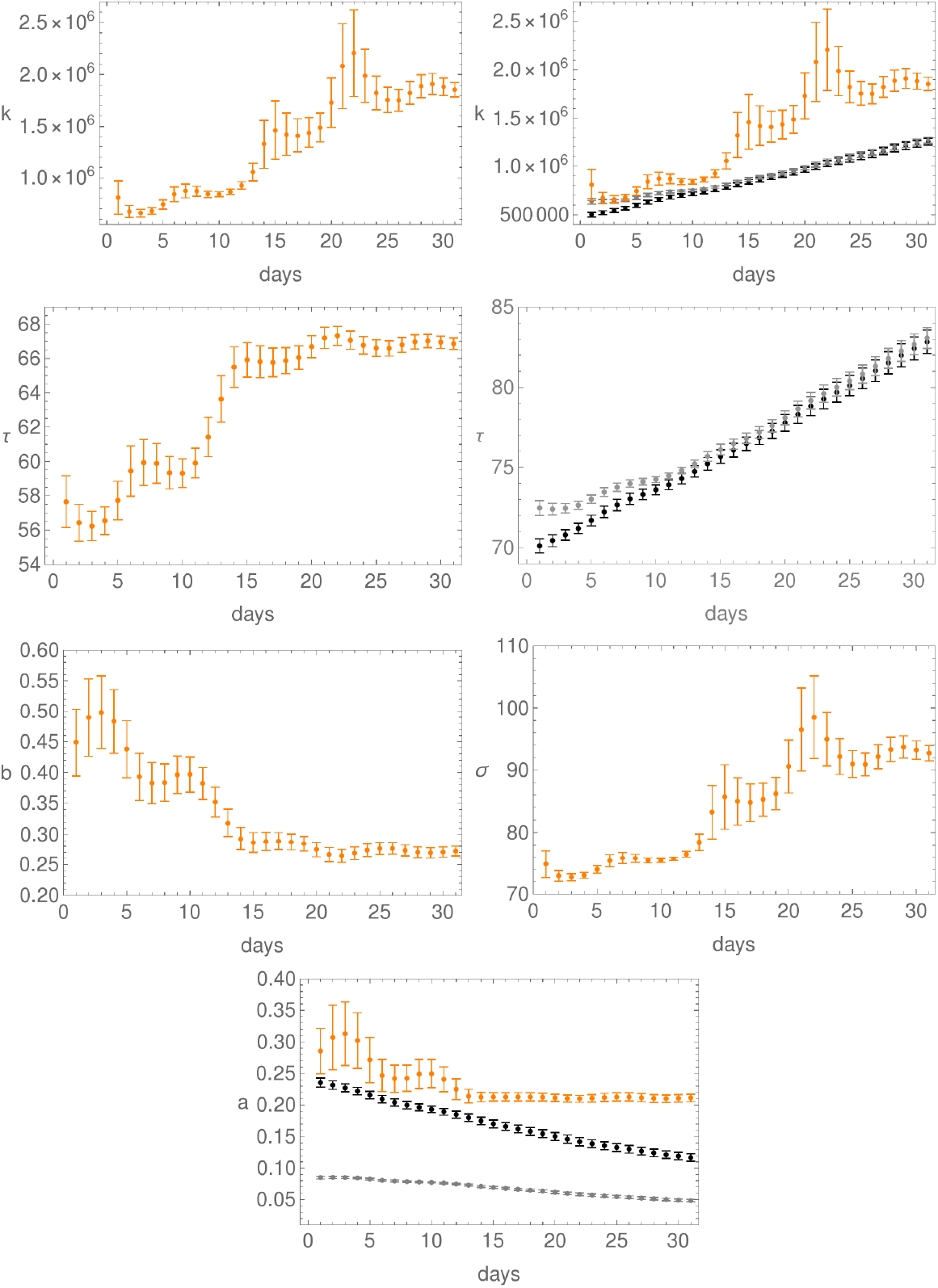
Evolution of the fit parameters in time, together with their confidence intervals, for US; day 1 is April 5. First row: parameter *k* for the double-logistic curve (left), and for all three curves (right). Here, as elsewhere, orange is for the double-logistic, black for the logistic, and grey for the Gauss-error function. Second row: the *τ*-parameter for the double-logistic curve (left) and for both the logistic and Gauss-error curves (right). Third row: plots of the parameters *b* (left) and *σ* (right) for the double-logistic curve. Last row: plot of the parameter *a* for all three curves.

**Figure A.29:**
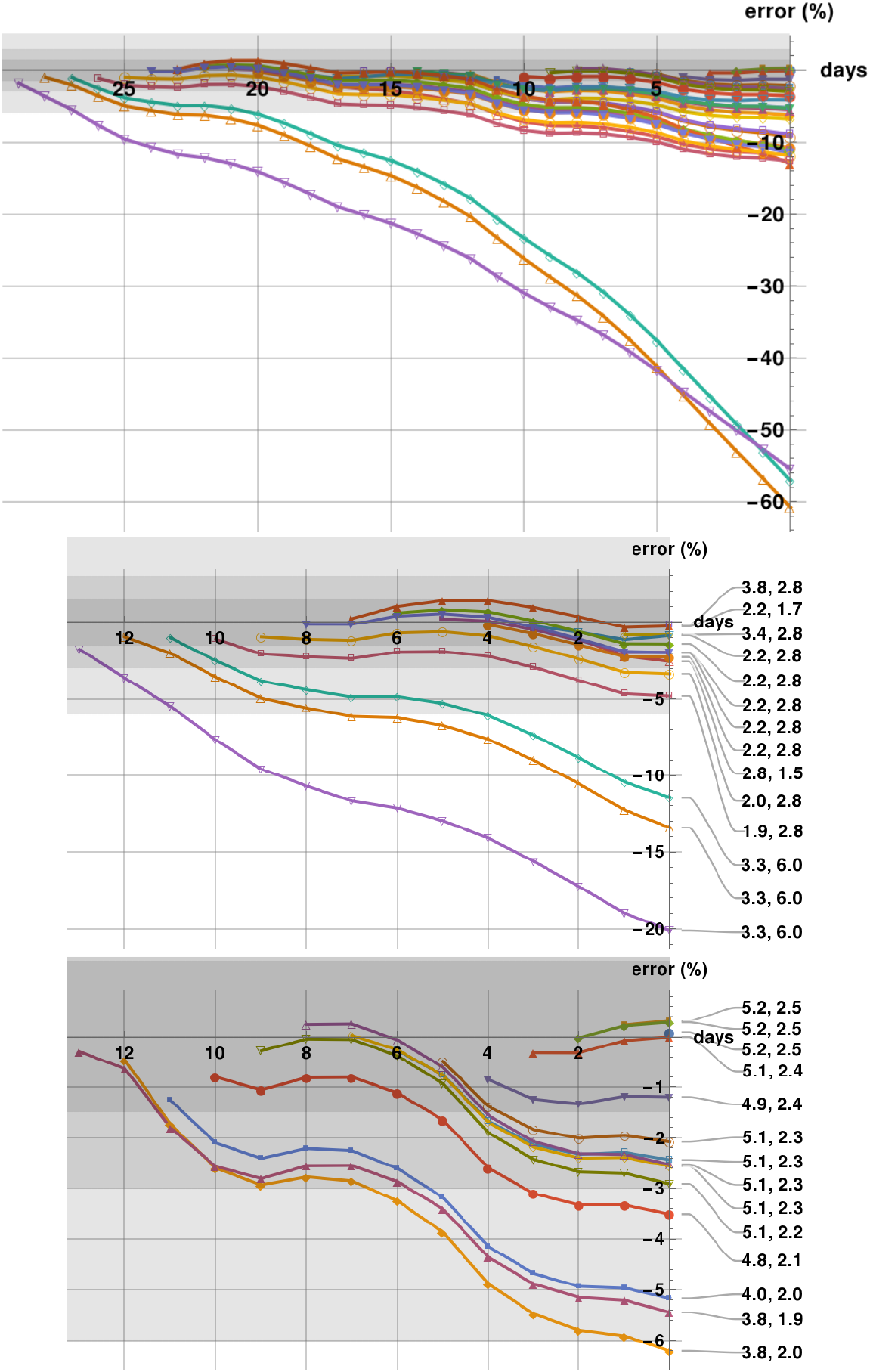
Predictability plots for US generated in the period April 5 - May 4. The first row shows all predictability strings with last prediction on May 5, which is the last day on the first and third rows. The shadings correspond to a 1.5%, 3%, and 6% error zones. The second row shows the predictability strings cut-off at April 20, with badness parameters indicated. The third row shows only these predictability strings which start within a two-weeks period ending on May 5.

**Figure A.30:**
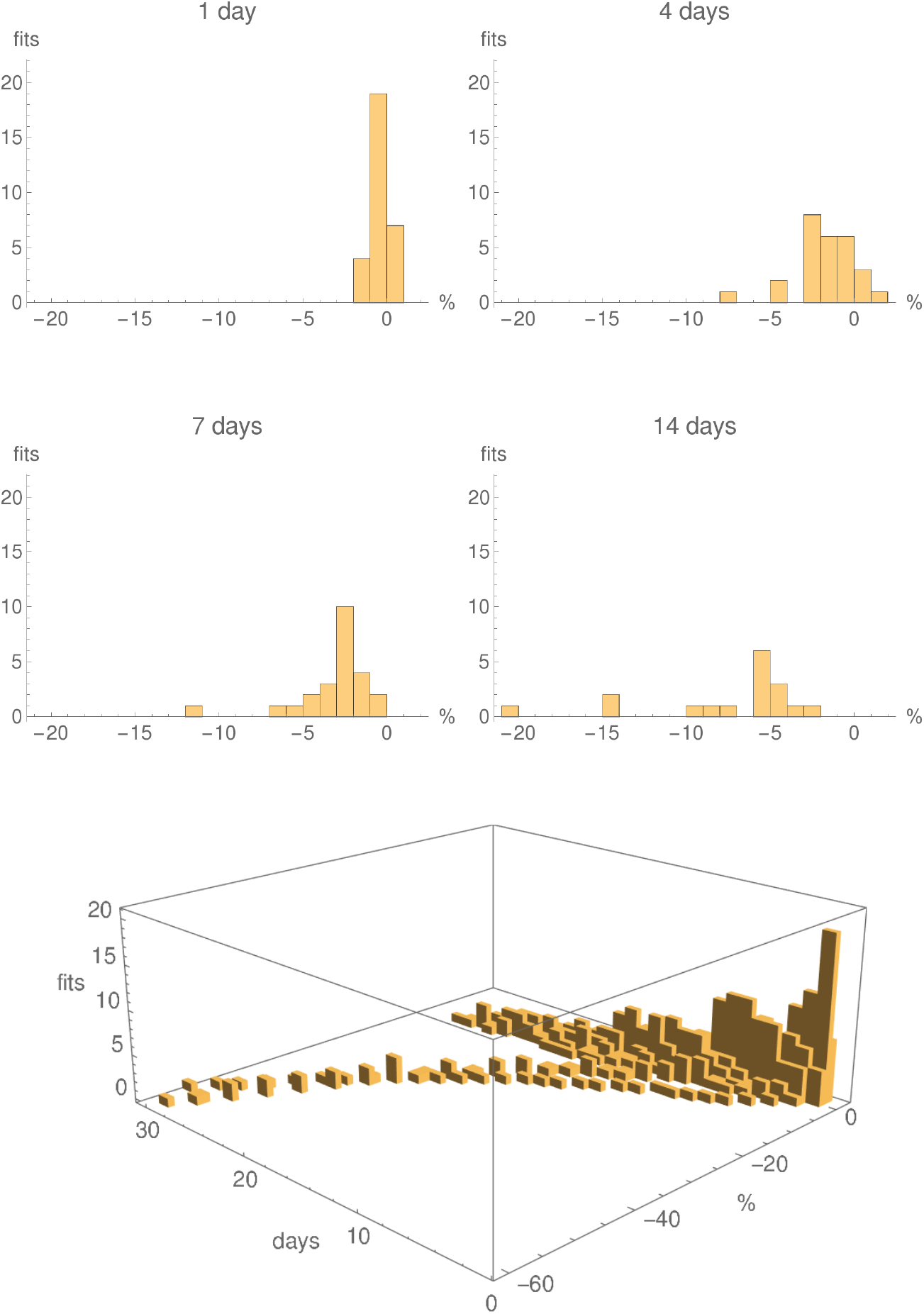
Histograms showing the numbers predictions with 1% error bins for the logistic fits with running parameters, for US, generated in the period April 5 - May 4, for 1 days, 4 days, one week and two weeks. The last row shows a 3D histogram for all 30 fits. The individual histograms in the first two rows are slices, at the given number of days, of the 3D histogram.

## B The Richards curve

A family of curves modeling growth phenomena has been proposed by Richards [2], and has been used by several authors. The Richards curve can be written as

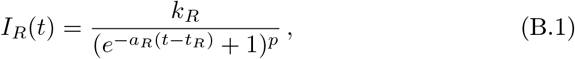

for some constants *k_R_* > 0, *a_R_* > 0, 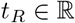 and *p >* 0. It holds that:

1. The function *I_R_* tends to *k_R_* as *t* tends to infinity, so that *k_R_* describes the total number of infected.
2. For large negative *t* the Richards functions behave as

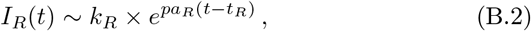 so that the initial rate of growth of the epidemics is *pa_R_*, with growth coefficient 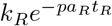.
3. For large positive *t* we have

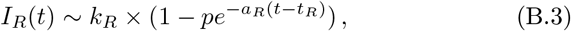

so that *a_R_* describes the dying-out rate of the epidemics, with decay amplitude 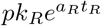.

We see that the Richards curve has the property, desired here, that the dying-out rate *a_R_* and the initial growth rate *pa_R_* are distinct. However, the Richards functions encode a relation between the initial growth coefficient, the final decay coefficient, and the rates just mentioned, which does not seem to be relevant for Covid-19. Our double-logistic curve (2.4) has one more parameter which prevents this correlation.

When *p* = 2, the double logistic curves (2.4) coincide with the Richards curves for specific choices of parameters.

## Footnotes

1 Note that this sign is opposite to that of the conventional definition of residuals.

